# The Milano Sforza Registers of the Dead: Health Policies in Italian Renaissance

**DOI:** 10.1101/2021.02.10.20249093

**Authors:** Elia Biganzoli, Ester Luconi, Patrizia Boracchi, Riccardo Nodari, Francesco Comandatore, Alfio Ferrara, Silvana Castaldi, Folco Vaglienti, Cristiana Panella, Massimo Galli

## Abstract

The institution of the *Liber Mortuorum* in Milan greatly anticipates death registrations in Europe. Introduced in 1450 by Duke Francesco Sforza for the early containment of plague it was daily compiled until 1801, reporting demographical data and causes of death, leaving a corpus of 366 volumes, an outstanding source for interdisciplinary research. Addressed to ascertain individual causes of death, it represents an unprecedented early example of disease monitoring and prevention. The paper discusses Sforza’s health policy in Milan, describes the Mortuorum registres features at the end of the 15^th^ century, and analyzes the distribution of the city deaths in 1480, a year without epidemic events. These data constituted the first entry of a database of the *Liber Mortuorum*.

## Introduction

This paper presents the *Liber Mortuorum* of Milan (henceforth named MiSfoRe) an exceptional source of sociological, anthropological, historical, and epidemiological data, put in place by Francesco I Sforza (1401-1466), duke of Milan from 1450 to his death, with the aim to timely track the occurrence of plague. As a seasoned soldier of fortune that he was, the Sforza had a pragmatic vision of a productive society that requested to keep people in good health, monitoring the health situation in the city and timely excluding the presence of plague in it. Thus, he imposed to register, since the late spring of 1450, all the deaths and ill people that occurred in Milan, with an accurate reporting in all the occurring individual cases of the demographical data and causes of the death. The Sforza’s will originated an enormous amount of registrations, collected during 350 years, which fill 366 manuscript volumes preserved in the Archive of State of Milan. This precious source of data has been scarcely utilized by scholars until the Seventies of the 20^th^ Century (Ferrario 1840; Cipolla 1974; Albini 1982; Zanetti 1976).

Differently from the Bills of Mortality of London, assembling quantitative data for statistical purposes, the qualitative approach of the Milanese way of death registration, with great accuracy in collecting individual data reflects unprecedented attention to healthcare issues, which will be confirmed by another Sforza, Francesco II, with the institution, in 1534, of the *Magistrato di Sanità* (health magistrate), a health office with various tasks, including that of continuing the assessment of the individual causes of death and the compilation of the registers.

To more effectively track the events of a plague outbreak, besides reporting, in the vast majority of cases, the parish and the *Sestiere* (the sixth part of the city) where each deceased person lived or died, the registers of the 15t^h^ century frequently included even more precise references on the place of death and information on the relatives or neighbors of the dead person. Such an accurate method of survey was maintained for more than one century, and strongly contributed to confronting the virulent plague epidemics which affected the city in 1476-78 and 1483-85 (Alter and Carmichael 1999; Vaglienti 2013).^1^

In this paper, after describing the health policy of the Sforza and the main characteristics of the city at the end of the 15th century, we depict the contents and the methods of the compilation of the MiSfoRe in comparison with the other main registers of dead in Italy and Europe. We also analyzed the aggregated causes of death in 1480, a year without plague, taken as an example of the causes and modalities of dying in the absence of epidemic events and more generally of the health situation of a large Italian city in the Renaissance. Information issued by the MiSfoRe has been used to build up a database, which is described in this paper, that will contain data about other years. This database contains, for now, only the data of 1480.

### The corpus of Mortuorum Libri

The State Archives of Milan presently holds a continuous series of 366 handwritten Registers (an example of a page of the Register of 1452 is shown in Figure 1), with only some discontinuities in the 15^th^ and early 16^th^ centuries. These gaps in the record were caused by a fire on the night between December 31, 1501, and January 1, 1502, at the headquarters of the Office of Health (*Ufficio di Sanità*), where the volumes were stored at that time. The series of registers from 1452 to 1801 is preserved and available for consultation, with some losses, mainly due to the 1502’s fire.

**Fig. 1.**
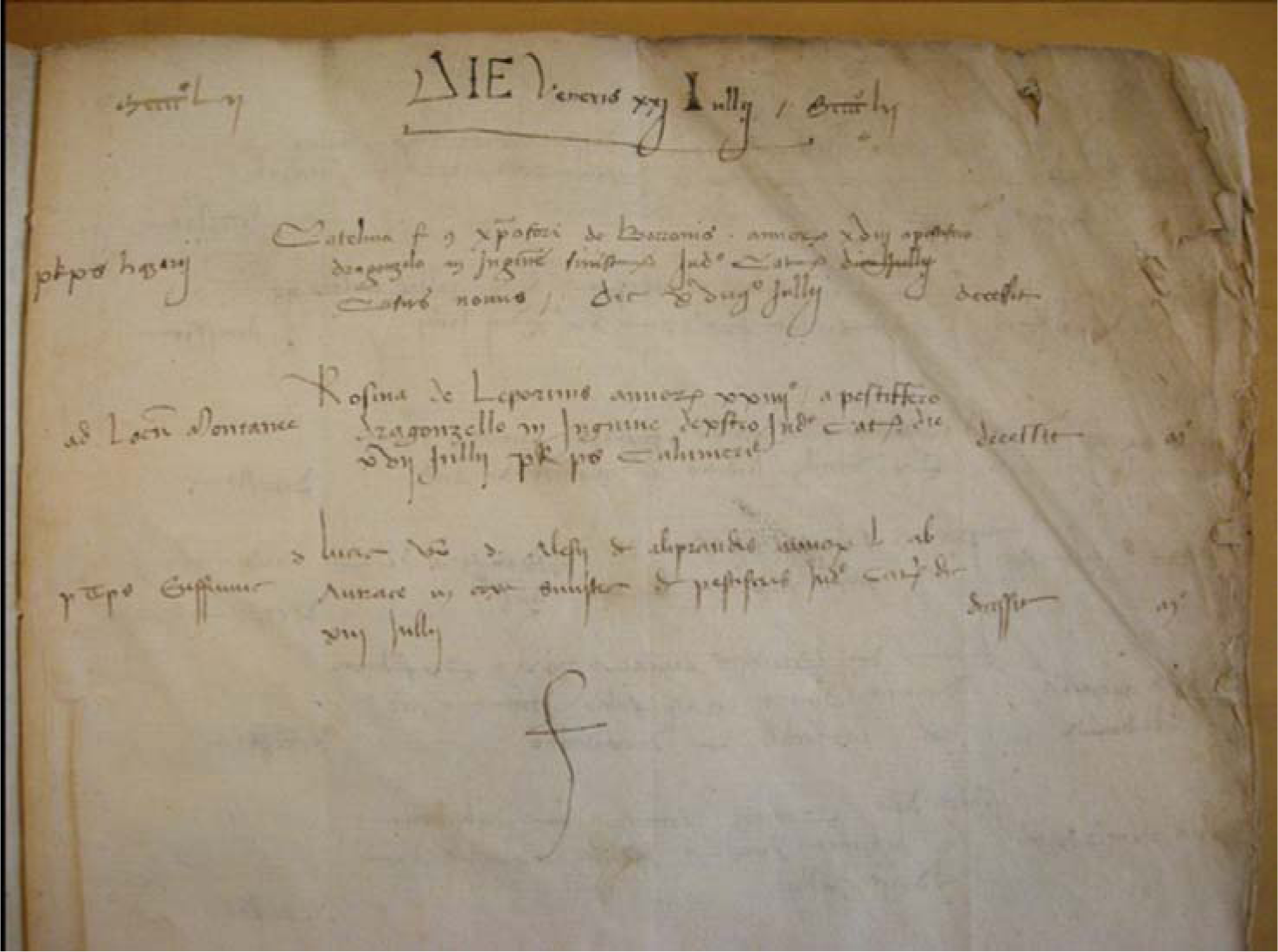
Sample picture of *Mortuorum Libri* register of 1452 (kindly authorized by State Archives of Milan). The transcription of the text, including the abbreviations used, and its translation from latin is shown below. “MCCCC°LII Die Veneris XXI Iullii MCCCC°LII 1457 IN THE VENUS’ DAY [FRIDAY], 21 JULY 1457

#### P R, p S Nazarii

*Catelina, filia quondam Christophori de Borronis, annorum XVIII, a pestiero dragonzelo in inguine sinistro, iuditio Catelani, die Iullii [*canceled]*, casus novus, die XVIIII° Iullii decessit* (*Porta* Romana, parish of *San Nazaro* (*in Brolo*).

(Catelina, daughter of the deceased Cristoforo de Borronis, 18 years old, due to a pestiferous bubo in the left groin, to the judgement of *Catelano* [the health officer in charge], [erased by the redactor of the registry], new case, died the day 19 July).

#### Ad Locum Montanee

Rosina de Leporinis, annorum XXIIII°, a pestiffero dragonzello in inguine dexstro, iuditio Catelanii, die XVII Iullii, Porta Romana, parochia Sancti Calimeri, decessit. m° (Montanee location. [open-air Lazareth] Rosina de Leporinis, 24 years old, due to to a pestiferous bubo in right groin, judgement of Catelano, parish of San Calimero, died). Dead

#### PT, PS Eufemiae

Domina Lucia, uxor domini Alesii de Aliprandis, annorum L, ab antrace in coxa sinistra de pestiferis, iuditio Catelani, die XIII Iullii, decessit. m°. (Porta Ticinese, parish of Sant’Eufemia. Lady Lucia, wife of Don Alessio Aliprandi, 50 years old, due to anthrax at left thigh of pestiferous nature, judgement of Catelano, the day of 13 July died. Dead.

*Finis* (the end of the day registration)”

The Registers of the 15^th^ century were written in Latin ^2^. Each record reported the day and the month in which the enlisted death occurred (Vaglienti 2013) and, for each case:

1. to which of the six *Porte* the death should be attributed.
2. the parish where the deceased person lived or died or would be buried.
3. the place where the death occurred if it did not correspond to the home of the dead person (the street, a hospital, an inn, a prison).
4. the name, surname, or nickname of the deceased person. For children, adolescents, and unmarried women, declarants reported (with few exceptions) the name of the father (specifying if still alive or deceased) or, if the father was unknown, of their mother. The foundlings were named *hospitalis filius* or *filia* (son, or daughter, of the hospital) or received a surname, most frequently *da Milano* or *Colombo*, with modifications in the frequency of their use and their variants over the centuries. The name of married women is often accompanied by the name and the surname of their husbands. Gender is not mentioned, but with a few missing cases, it is easily inferable from the person’s name or adjectives used.
5. the social and professional status of the deceased (of the father in the case of children or the husband in the case of adult women) including titles indicating nobility, ecclesiastical or professional status, or titles deriving from military or administrative functions as well as the occupation of servant, merchant, craftsman or worker, the condition of poverty, or the status of a prison inmate.^3^
6. the age at death, expressed in years for adults, and in months, days, or even hours for infants.
7. the cause of death. Simply reported as plague or other than plague in the first registers, the causes of death became shortly thereafter accurately described, in some cases with a level of detail such as that provided in the following sample, taken from the register of 1459: “Long-suffering of quartan fever, the night before he had a paroxysm, aggravated by an indigestion that caused heart palpitations and blurred intellect, then evolved into syncope and apoplexy that led to the death ”. The information about the the duration of the illness leading to death is usually collected because it is an important indicator of the presence of plague which was believed to lead to death within three to four days.
8. the name of the official in charge of inspecting the bodies and determining the causes of death which will then be transcribed on the register along with all other information requested. This official could be a ‘collegiate’ doctor (indicated in this case with personal name and title) or a health official appointed to this specific duty; less frequently, the Elder of the parish or, in exceptional cases, the gravedigger.

### The MiSfoRe and the others: registering mortality and morbidity in Italy and in Europe

As established in 1450, the 15^th^-century portion of the Liber Mortuorum death registers, the MiSfoRe, are undoubtedly older than the most renowned source of morbidity and mortality data of the European modern age, the Bills of Mortality of London. The first London Bill of Mortality is believed to date from November 1532. Produced intermittently in several parishes of London during outbreaks of plague, where parsons reported the total number of plague deaths and bodies buried each week, the Bills compiled by Worshipful Company of Parish Clerks the from 1611, this duty was imposed by James I (Slauter 2011). Beginning in 1560, pairs of older women in each parish were charged with examining the dead and determining the cause of death. This choice of relying on the practical experience of older women and their knowledge of the local social context was considered adequate for the quantitative approach to recording the deaths of the Bills (Boyce 2020). The clerk of the Parish Company had the task of sending a copy to the Privy Council within the following day (Heitman 2020). The London Bills only began reporting causes of death and the gender of deceased persons in 1629, while the age at death was reported from 1728 onward (Mazur 2016).

Thus, the MiSfoRe pre-dated the London Bills of Mortality as a reliable and stable death register by more than a century. Moreover, the scope of the MiSfoRe exceeds that of the London Bills of Mortality: from the introduction of this register in 1450, the MiSfoRe focus on every single case of death, providing information on the age and sex of the deceased, the social condition, and the cause of death. As a result, the MiSfoRe represent a much richer source of information. Furthermore, compared to the London Bills of Mortality, the MiSfoRe were the manifestation of a direct government intervention, mainly based on specific dedicated health officials, tasked with issuing judgments on the causes of death of each individual and registering every death daily. In contrast, the Bills reported weekly and collectively the burial dates of deceased persons whose causes of death were evaluated by elderly parish women (Rollo-Koster 2016). However, before the plague of 1592 the London reports were available only to the Privy Council, the sovereign, and London’s Lord Mayor, thereafter the Worshipful Company of Parish Clerks began to produce a large-scale broadsheet and post them in public areas. As a consequence, the Bills of Mortality of London became the most renowned source of morbidity and mortality data, and form the primary source of information for public health sciences since its early origins (Heitman 2020). The identification by John Graunt, as early as 1661, of some fundamental demographic laws such as the numerical regularity of births and deaths, the relationship between the sexes at birth and death, the percentages of each cause of death relative to the total of deaths, and the consequent predictability of many biological phenomena, derives from the opportunity provided by access to the data of the Bills of Mortality (Sutherland 1963).

On the contrary, the MiSfoRe have remained manuscripts until 1774 - three years after the introduction of systematic surveys of the Lombard population commissioned by the government of Maria Theresa of Austria (Zanetti 1976) - when they began to be published in print, but their use remained limited to Health Offices. Scholars only began to be interested in the MiSfoRe registers in the 1830s (Ferrario 1840) and incredibly this extraordinarily rich source of data remained unused until the 1970s (Cipolla 1974; Albini 1982; Zanetti 1976).

The other death registers of the main Italian cities experienced a similar fate. For example, in Venice, from 1504, the parsons were instructed to make daily visits to sick people living in their parish and record in a book the information about the illness and if a doctor was taking care of the case. Daily information collected by parsons of Venice was transmitted to the *Ufficio della Sanità* (the Office of Health) and, from 1537 to 1805, systematically reported in the *Necrologi* (necrologies). From 1537 to 1565 the registered data included for adult men the name, for others a more generic definition such as “a child” or “a widow”, the occupation (for adult men, for others, when provided, the one of the father), and the parish. From 1565 the included data become more similar to those reported in the Milan registers, including name, surname, age, length and nature of the illness, occupation, gender, and parish. From 1731, the physicians who visited the sick people had to certify the nature of the illness, and their names were reported in the registers, but the clergy remains responsible for the transmission of the information (Bamji 2016).

The registers of Venice was used to make a digital reconstruction of the plague outbreak in Venice during 1630-1631: in particular, they presents heatmaps regarding the number of death in parishes along time to verify if the peaks occurred in synchrony between the different parishes and they used data to apply epidemiological models to better understand the modality of the contagion. The limitation of this work was that not all the records have been preserved (Lazzari et al. 2020). Another work about this plague period focused on the period of three days during the peak, including the sex ratio and the distribution of age of death for plague and other causes. They also investigated the fact that plague could have an impact on the other causes of death, and finally, they described the spatial characteristics of the epidemic. The limits of this work were the fact that there are some “dubious cases” about if the causes of death were plague or not and the records were compiled by many people, so the descriptions of the same symptom can be varied (Ell 1989).

The *Libri neri dei morti* of Florence established in 1385 by the Board of the *Grascia*, an agency to regulate the provisioning of the city, are the oldest example of death registers in Italy. Compiled up to 1778, they contain the name, the paternity, the date of the death and the place of the burial, and sometimes the age and the social status of the deceased person. From the early 17^th^, the occupation was added. A second register was provided by the Guild of Physicians and Apothecaries and covered the period between 1450-1808. The gravediggers, who belonged to the Guild, were the source of information, reporting the names of those they buried and the date of the death. From 1780, parishes got the task to report information about the deceased people (Parenti 1943; Cipolla 1978). Apart from rare exceptions, there are no records of infants and children deaths. Therefore, even if the Florentine registers were established before the MiSfoRe, the data contained in them are generally less complete. Moreover, both in Venice and Florence the registration of the dead was not a responsibility directly assumed by the state government.

Diana’s work on the Florence Registers explores the correlation between profession categories and intestinal infections. The investigation concerned the plague periods 1424-1425 and 1630. Nevertheless, the registers present some omissions in recording professions and also for deaths for some months during the year, for example, in 1425, records were present from the first day of July to April 18 (Diana 2002).

In Italy, many other registers have been established to detect the presence of the plague in the city and suburbs. In Mantua, for example, from 1456 there were lists of death and indications about the causes of death, and in 1839 Count Carlo D’Arco used the Mantua registrations to produce statistical observations of the fluctuations of causes of death during the year and the change in the main causes of death in the various historical periods (Zanetti 1976). In 1501 there were some gaps, especially in periods with the plague; the completeness is estimated to be 2/3 of the actual number of deaths. From 1502 there was more regular monitoring (Romani 2016).

Verona’s registers established from 1629, were also similar to those of Milan: dead people were reported day by day (women, men, and children), along with the cause of death and socio-demographic information such as age, country, and duration of the disease.

Outside Italy, Barcelona’s registers began in 1457 (Zanetti 1976). The responsibility for compiling the Barcelona registers was entrusted to the letter carrier, who had the task of obtaining and reporting daily information on the number of funerals and baptisms in each parish. It is uncertain whether these records also included infants (Smith 1936). The Barcelona death registers were constituted by the city council and were used to detect the presence of the plague.

In Smith (Smith 1936) concerning the seasonality of the peak of plague period in different years and other causes of death in periods without plague, it has been remarked that possibly there was an underestimation of the other causes of death, and infant and birth deaths are not reported. Moreover, the data are discontinuous for many months (but they are complete for at least a quarter of the year) and those about baptisms are extremely discontinuous.

### Milan in late 15^th^ century: the the social and demographic context

We now present the population and organization of the city to give an idea about the importance of the city of Milan in the considered period; the MiSfoRe reflect such importance through indications about the parishes and the *Sestieri* where the death events occurred, for which we made epidemiological analysis, as presented below, taking in consideration the *Contrade* that had a connection with the two geographical indications. This section is important also for the database, as described later, because geographical information is also contained in it.

In the absence of fiscal records and a land register, it is more difficult to estimate the population of Milan than in other cities of medieval Italy having those documents (Albini 2021).

In such a context, it can be understood the extraordinary value assumed, starting from the second half of the fifteenth century, has an extraordinary value (Vaglienti 2013). In 1288, at the time of Bonvesin de la Riva (a Lombard author, probably Milanese) it was estimated that Milan could have 200.000 inhabitants (Bonvesin 2009; Albini 2021). However, in the first decades of the 15th century, due to the recurrence of epidemics and the considerable migratory flows from the city dictated by the growing economic difficulties of an increasing number of unemployed and indebted wage earners, Milan and its territory were much reduced compared to that of 13^th^ century.

From 1450 Sforza’s policy and the internal security guaranteed by the new sovereign led to a demographical increase. There was offered a large number of new citizenships, favoring the immigration; in particular, there was immigration not only of the but also of the workforce from the countryside, from the other parts of the Milanese dominion, and other states. In 1480 Giovanni Ridolfi (Florentine politician and diplomat) was so impressed by the dynamism of the Milanese population that he dramatically overestimated its size to 800.000 inhabitants. More realistically, the stable population, excluding the fluctuant one (the people temporarily entering the city for work and temporary residents) (Albini 2021), probably did not exceed 100,000 units (Chandler 1987).

Considering a city as an urban agglomerate with at least 10.000 inhabitants (De Vries 2013), in the 16^th^ century, Italy had the largest urban population in Europe ^4^. At the end of the 15^th^ century, Milan was one of the major European centers for the production of arms, armor, and fabrics in Europe and a trading center of primary importance, connecting northern Europe with Italy (Tucci 2011).

Milan’s map reflected the expansion of the city over time: Roman walls, built between the first century B.C. and the end of the third century A.C., delimited the development of the ancient *Mediolanum* (in the middle of the plain) during the Roman period and were largely destroyed thereafter. A second wall ring was built between 1156 and 1171 and included the medieval city. The building of the most recent wall ring began in 1459 with Francesco Sforza and was completed in 1560, during the Spanish domination. Of all three circles of walls, only a few remains are now visible, incorporated into the further growth of the city in modern times. The medieval city was divided into six parts, called *Sestieri*, each of which drew on the map of the city an irregular triangle, whose vertex was the *Palazzo della Ragione*. The *Palazzo della Ragione* for centuries played the role of *Broletto* and was located in *Piazza dei Mercanti*, in the center of the city.

The base of each triangle was along the medieval ring of the city walls. Each *Sestiere* was named after the corresponding *Porta* of the city - as originally the population of each of them was responsible for the defense of the corresponding gate on the city walls (Tucci, Ronza, and Giordano 2011). Each *Sestiere* was subdivided into five *Contrade*, which were contained in the second (medieval) ring of walls (Fig. 2). One of them, not necessarily the *Contrada* closest to the city center, but probably the richest one, bore in each *Sestiere* the denomination of “noble” (Milano Città Stato, 2020).

**Fig. 2.**
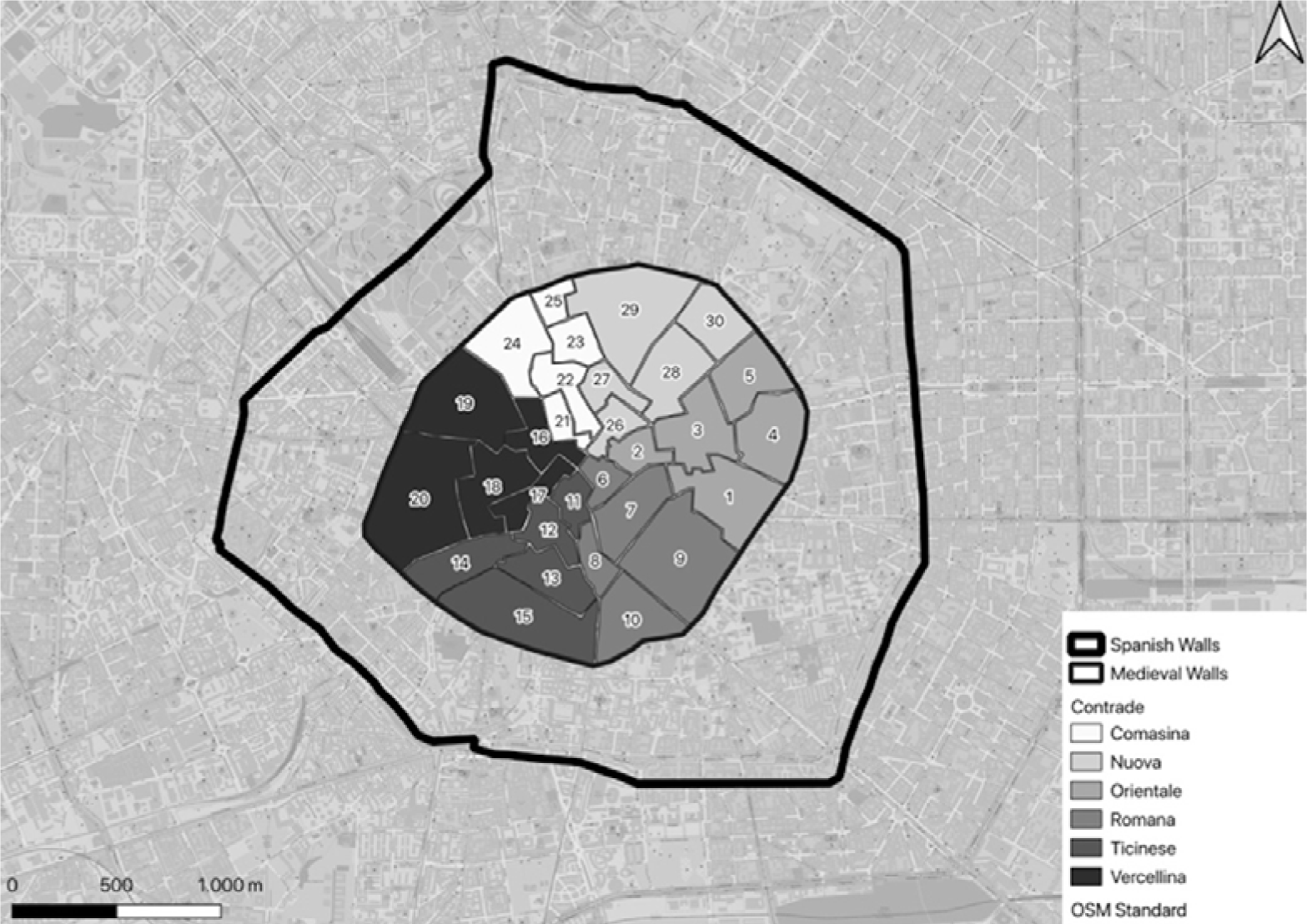
The position of the *Contrade* in the city of Milan along with the distribution in the *Sestieri*. The position of the medieval walls and the Spanish walls are also represented. Digitalization and reconstruction of the material found on internet MacMoreno, Sestieri e Contrade di Milano, the Creative Commons Attribution-Share Alike 4.0 International, https://en.wikipedia.org/wiki/GNU_Free_Documentation_License)

The *Contrade*, as well as the *Sestieri*, no longer had any political or administrative function at the end of the 15th century. However, the distribution of deaths in the *Contrade* in a year without plague or other epidemics can offer an indication of the populousness of the various parts of the center of Milan and a comparison with the parts of the city that arose outside the circle of medieval walls.

The city doors were called, going clockwise starting from the North, *Porta Nuova, Porta Orientale, Porta Romana, Porta Ticinese, Porta Vercellina,* and *Porta Comasina*. Originally located on the second ring of walls, each door of the city was transferred thereafter to the new wall. The part of the city included between the second and third wall was divided among the *Porte*, but not attributed to the *Contrade*. Table 1 reports the names of the *Contrade*, their surface in square meters, the *Sestiere* they belong to, and the number of parishes located in each of them along with the name of the parishes.

**Table 1.**
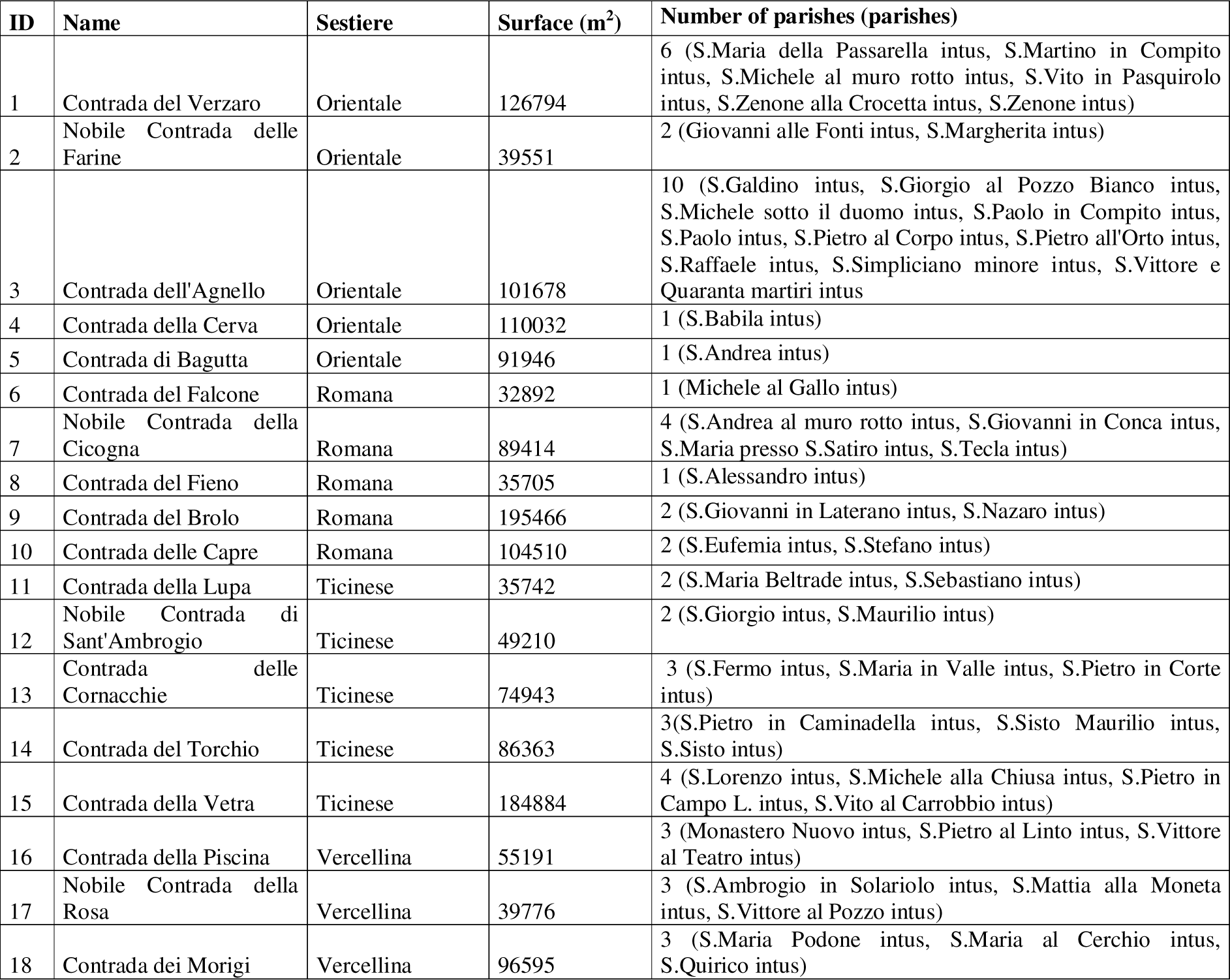
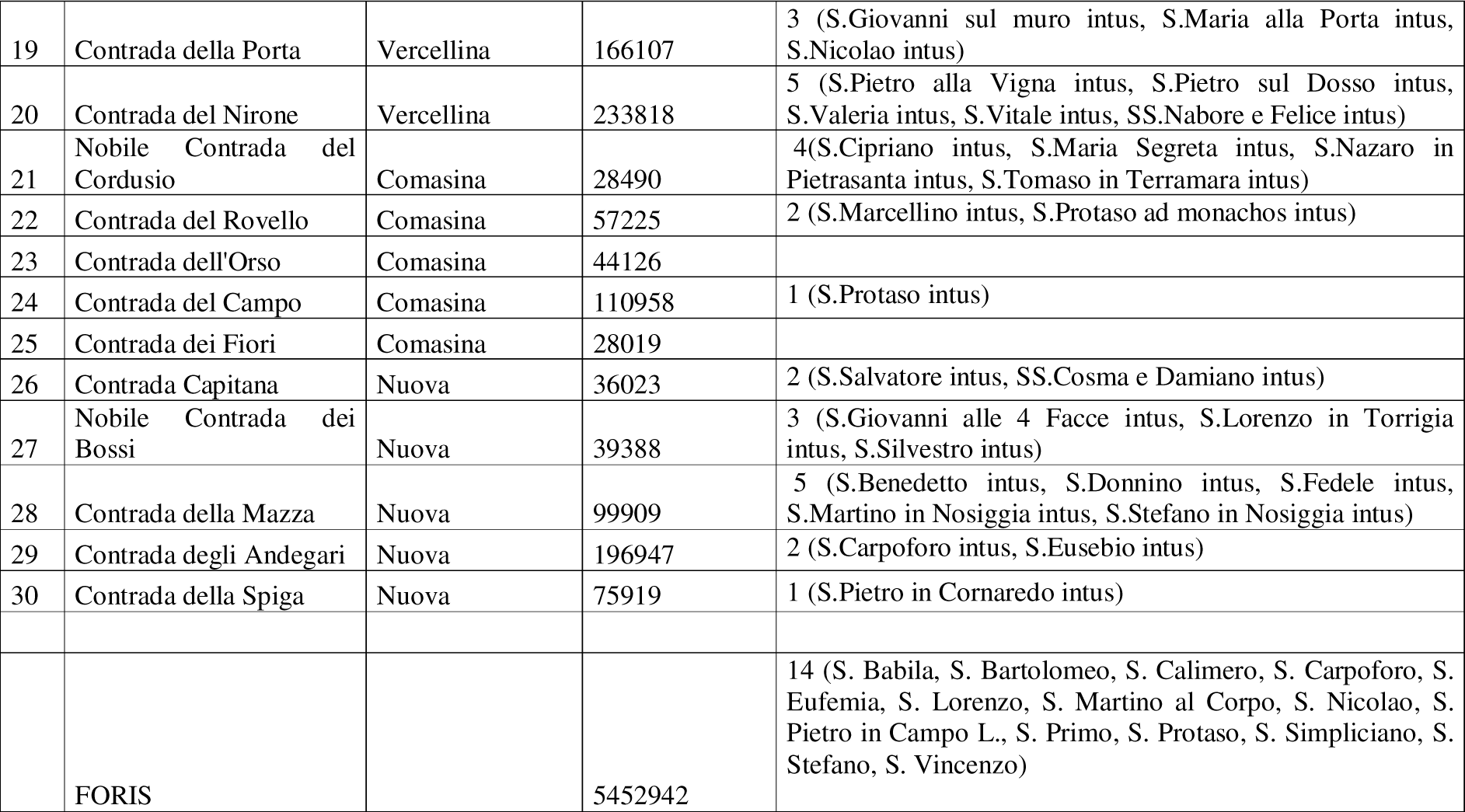
The Contrade of Milan, their surface, the Sestiere to which they belong, and the number of parishes whose reference church was located in the territory of each of them. The names of the parishes in each Contrada are also presented.

This division in *Porte* did not reflect the social organization of the city in the second half of the 15^th^ century when it was mainly based on the parishes (Colombo 1955). In the mid of 15^th^ century, a relevant part of the parishioners of many large parishes located inside the medieval walls, lived outside the wall ring and the territory of a *Contrada*.

Between 1400 and 1550 the area between the medieval walls and the Spanish walls was characterized by different activities and territory uses, including areas dedicated to periurban agriculture, the so-called *Cassine* (farmhouses). Firstly these properties were ecclesiastical, but in fact, managed by lay farmers who later became the owners. Artisan workshops (for moleskine and silk as examples) and activities that could harm public health (such as fabric dyeing workshops); were also placed in these areas outside the medieval ring of walls. In general, the activities in these areas represented the expansion outside the medieval walls of those of the nearest part of the corresponding *Sestiere* in the medieval city. However, in 1412, there were outside the medieval walls some heavily populated areas, mainly inhabited by lower-class people, which were interposed among the cultivated plots. Some areas outside the center of the city were also chosen as a place of residence by upper-class people and some nobles. (Boucheron 2006).

In the Middle Ages and Renaissance the different parts of the city, at the neighborhood level, were characterized by a certain social heterogeneity, even if nobles more often preferred to stay in the peripheral area because the center was characterized by commercial activities (Folin 2009).

Originally established on criteria not based on administrative needs and in any case before the subdivision of the city for defensive and military purposes, the area of competence of each parish could include, particularly in the city center, parts of the city belonging to two or more different Porte or Contrade. As regards the distribution of the different social strata, the central part of the city was not reserved for wealthy people and nobles. Nevertheless, based on the economic activities carried out and the social composition of the inhabitants four concentric rings could be somewhere defined. The central one included the majority of the shops and homes of merchants and retailers. The second was mainly a residential area with the houses of noble and wealthy people. In the third ring, there were many craft workshops, especially those producing textiles, weapons, armor, and trade places and stores near waterways, artificial canals called *Navigli*. The last area towards the periphery and outside the second ring of the walls was populated by workers and the poorest part of the citizens, even if some noble houses were also present. Churches and monasteries were scattered throughout the city but particularly concentrated inside the line of the medieval walls. Even if areas that could be considered completely homogeneous as regards the social composition were virtually absent, some parishes were predominantly inhabited by noble and wealthy people, and some streets and neighborhoods were inhabited by artisans and traders with similar activities called *Mestieri*. In particular, there was a radial distribution of commercial activities along the roads that connected the gates to the city center, while residential districts were located between these trade routes (D’Amico 1994).

### Discover the plague soon, inspecting every single death

Milan was spared by the Black Plague of 1348, but since that year, the Milanese domain started to activate early a monitoring and control system on the diffusion of plague and other epidemics (Pasi Testa 1976). In particular, from 1374 the lords of Milan order all mayors that, through the presbyters, the parishioners, the neighbors, or the elder of the parishes, it should be verified, on a daily basis and with the indication of the time, when and where cases of plague might have emerged among the observed deaths.

In 1399 the first duke of Milan Gian Galeazzo Visconti, to face plague epidemics entrusted his general vicar as “*commissaries specialiter* […] *pro conservation sanitas ducatus Mediolani*” (i.e. special commissionaire for the health conservation in the Milanese domain). Among the functions to be assumed by the new government apparatuses in the entire domain, there was the obligation to draw up lists of the deaths with an indication of their causes.

Almost forthy years after, in 1438 the third duke, Filippo Maria Visconti, ordered that health professionals (doctors, surgeons, barbers, and apothecaries) should enlist the names of the sick person they were caring for while the *Anziano* of the parish should notify all the sick and dead people by a maximum of five days from the event (Albini 1982).

At the beginning of the century, Duke Gian Galeazzo Visconti had already charged the *Anziani* of the parishes to report suspected cases of plague (Rollo-Koster 2016), including those that occurred among the members of the clergy (Cipolla 1974) (Zanetti 1976). The role of *Anziano* (Elder), a layperson elected by the parishioners, represented a reference for the population of the neighborhood. They knew all their parishioner and they had the task to indicate to the civil authorities people who needed help and support (poor and frailty people, foreigners) and potential criminals. They also had to monitor the environmental quality of the area under the jurisdiction, so they had to supervise who produced polluting waste or sewage (Antonielli 2008). Parish Elders had the task of assisting in the act of burying all the corpses in their parish, even those who would not receive a religious funeral for some reason, and collecting the burial procedure fee (Vaglienti 2021).

Francesco Sforza’s project takes the early detection of plague cases to a higher organizational level and accentuates the attention paid to individual conditions in each case of death. The task of compilation and conservation of the registers was assigned to the *Ufficio di Sanità* (Health Office), which was composed of a *Catelano*, a doctor surgeon specialized in detecting epidemic diseases, a barber, in charge of minor surgery, a carter, two burials, a notary, two cavalrymen, three servants, a messenger in charge of carrying the death reports and, at the top, by a commissioner and a variable number of deputies, who were entrusted of preserving public health not only in the city but of the entire domain (Cipolla 1976). The *Ufficio di Sanità* (Health Office) originally stood in a building in the *Camposanto* (very near to the Dome of Milan). Following a fire on the last night of the year 1501, the Office was then moved to a building behind the *Corte Vecchia dell’Arengo* (now the Archbishop’s Palace) (Vaglienti 2021). In times of epidemic, an expert doctor of plague able to promptly identify the signs of the contagious disease, as well as to treat infected people, was to be added to the Health Office (Vaglienti 2021). The chancellor-notary of the Health Office must receive the certification of suspected infirmity and death (natural, traumatic, violent) and should register it in the Books (Vaglienti 2020a) ^5^.

During Francesco Sforza’s government, the professionals who composed the Health Office should report directly to the Duke (Vaglienti 2020a).^6^

Two relevant figures not included in the Health Office who played an important function in the death registration system were *officiali delle bollette*, parish elders, the latter had indeed always been involved, as described above, the attending physician and the *Catelano.* Parish Elders had the task to visit the home of any dead person in the parish, collecting the data requested for the registers, which should be transmitted to the *Ufficio di Sanità*. In the 15^th^ century, each deceased person was then directly inspected by Catelano or by a doctor charged by the Health Office (Vaglienti 2013). First, the *Catelano*, reconstructed the clinical history of the patients, then he inspected the body of the deceased and in the presence of a cause of death that could lead to an epidemic, he consulted with his colleagues or interrogated the death register for similar cases and finally produced the diagnostic report which was finally delivered to the health office to compose the transcript of the event in the book of the dead (Vaglienti 2021). The attending physician could be questioned by the parish elders in the act of finding information on the deceased and issuing the death certificate that permitted the *sepeliatur* (literally: let them be buried) in the absence of which it was forbidden to bury anyone, regardless of social status. Consequently, people were obliged to report every death to the competent authorities. The *officiali delle bollette* had the task to control the non-resident population of Milan, notifying those who took accommodation in the accommodation facilities (Vaglienti 2021).

In 1534 the duke of Milan Francesco II Sforza, always to assess the potential presence of the plague in the city instituted the *Magistrato di Sanità* (health magistrate) that had the authority over the whole state over anyone who violates health orders or anyone who causes damage to the public health. The control of epidemics through this structure took place through prevention, control of access routes, fines or penalties for people who did not comply with health regulations, and the isolation of areas at risk of the city. The health magistrate also had the task of checking the usability of the houses and checking the work activities considered harmful to health. It also had to register the causes of death. The scribe had the task to write in the book of the dead the name and the information of the deaths of the days and he produced the *Sepeliatur*. The chancellor had to maintain four registers: in the first one, he described all processes and the complaints along with the cause, in the second one the decrees of the prefects, in the third one the information about the treasury and in the last one, which is compiled only in the cause of plague, the names of the buryes and those in charge of “sanitizing” the objects and furnishings that belonged to the plague sufferers (*purgatori)* and the names of the infected families together with the places where the aforementioned were sent to remove them from the city. There were also doctors who had the task to ascertain the cause of death in the event of sudden death or if a non-collegiate physician has dealt with it and the judgment was uncertain.

The commissioners of health had the task to maintain environmental health conditions in the city. There were also external commissioners who were activated in case of plague outside the city and were in charge of enforcing and imparting the rules aimed at containing the contagion (Lampugnani 1578). The health magistrate remained organized in those terms until 1749 when Empress Maria Theresa led to the reduction of the number of quaestors to one. Finally, in 1786 the Austrian government decreed the abolition of the Magistrate (Visconti 1911).

Fighting the plague in early modern Italy meant putting in place measures of containment, mitigation, and quarantine. Prevention consisted, in the first place, in preventing the plague from reaching the city, for example by not letting infected people in and by interrupting trade with the places where there was the disease. If the plague entered the city, containment was implemented by closing the houses with the sick, imposing a lockdown, and isolating the affected areas of the city. Mitigation consisted of contact tracing and all the measures to prevent crowdings, such as the closure of churches and the cessation of some work activities. Quarantine took place in various cities in the Lazarests, such as Milan and Venice, this was done to move away infected people from the city (Henderson 2020). Examples of those measures could be seen in the duchy of Milan during the plague of 1576-1577 (also known as San Borromeo’s plague). From 1575 there was a prohibition of entry into the city of things and people from the territory in which there was the plague (Trento, Verona, and Switzerland), in 8 April 1576 the prohibition was for Mantova and in subsequent months also for Venezia and Sicily. On 10 April there was the suspension of the markets and on 12 April there was a prohibition to enter the Milanese duchy without the *bolletta di sanità* which certifies the place of origin of the person and the goods. In the same period, there was a prohibition to exit the city for the categories of people very important to the economy and the public function of the city.

In August 1576 the plague enter the city. From the first of October was instituted a quarantine for females of all ages and social status and children under 8 years. On 29 October 1576 started the quarantine for all the population in the city and the population living in a ducal territory in a presence of plague cases. The quarantine was maintained, with various concessions according to the epidemic trend. From 21 June 1577, the provision waned. On 20 January 1578, the city was declared free from plague (Bianchi Riva 2020).

Moreover, the individual responsibility of health officials is emphasized and moral integrity would be warranted through personal commitment and legal liability. The officials engaged in registering the deaths in the MiSfoRe were personally responsible to ascertain the causes and had to personally respond to a possible investigation by the competent judiciary where irregularities are suspected.

In most cases, the Registers report not only symptoms and supposed causes of sicknesses causing the death, but also the elapsed time from the beginning of symptoms and the death or the visit of the health officer, and rarely the possible presence of symptoms in the relatives living in the same house. Identifying the course of the disease along with the duration of the illness was important to be able to exclude the plague as the cause of death. As a result, the health officials were required to compile a daily list of seriously ill patients. The occurrence of death after the fourth day of disease was considered a sufficient proof to exclude the suspicion of plague (Cohn Samuel K and Alfani 2007) The scrupulousness collection of information requested by Sforza required that official agents visit sick people, including the poorest, even during plague epidemics, in whatever condition they were in. Health officials were in charge of inspecting the corpses personally, collecting the deceased’s data, and registering them on the same day or within a day after death. The same duty was imposed on the graduate doctors (those who have attended medical faculties) for their patients who died at home. Although aimed, at the time of their establishment, at the rapid detection of plague, the MiSfoRe began in a short time to report also the modalities of the violent deaths, including those caused by accidents at work. It suggests that the ducal government intended to collect information to assess the risks associated with certain works or environments (Vaglienti 2020a) and finally the details of any cause of death.

The collection of community-level information as well as concomitant pathologies and modalities of violent deaths indicates that the death registration program associated with MiSfoRe implied a foresightful vision of the community as an interdependent whole, beyond class and economic distinction. As a result, the deaths of secular and ordinary clergy who died outside convents and religious institutions were registered, although deaths that occurred in monasteries were not registered.

### The Database project

The interdisciplinary approach to the Registers has implied a proposal of a database to make accessible the information contained in MiSfoRe to scholars of different disciplines (e.g. historians, medical historians, anthropologists, clinicians, epidemiologists).

The database is a project of *Università degli Studi di Milano* in collaboration with the Direction of the State Archives of Milan, which is currently not funded. It will contain all the data of plague years and two years between plague period, after that the inclusion of some periods judged to be of particular interest from a historical and/or epidemiological point of view. Data are entered by a manual transcription of the information contained in the registers including, when available, information about household organization, professional positions, and social status of the deceased persons. Thus, the project implies for each domain (Figure 3) as well as information available in the Registers as external links toward literature and archival sources to enrich the social landscape which molded the health conditions of the population.

**Fig. 3.**
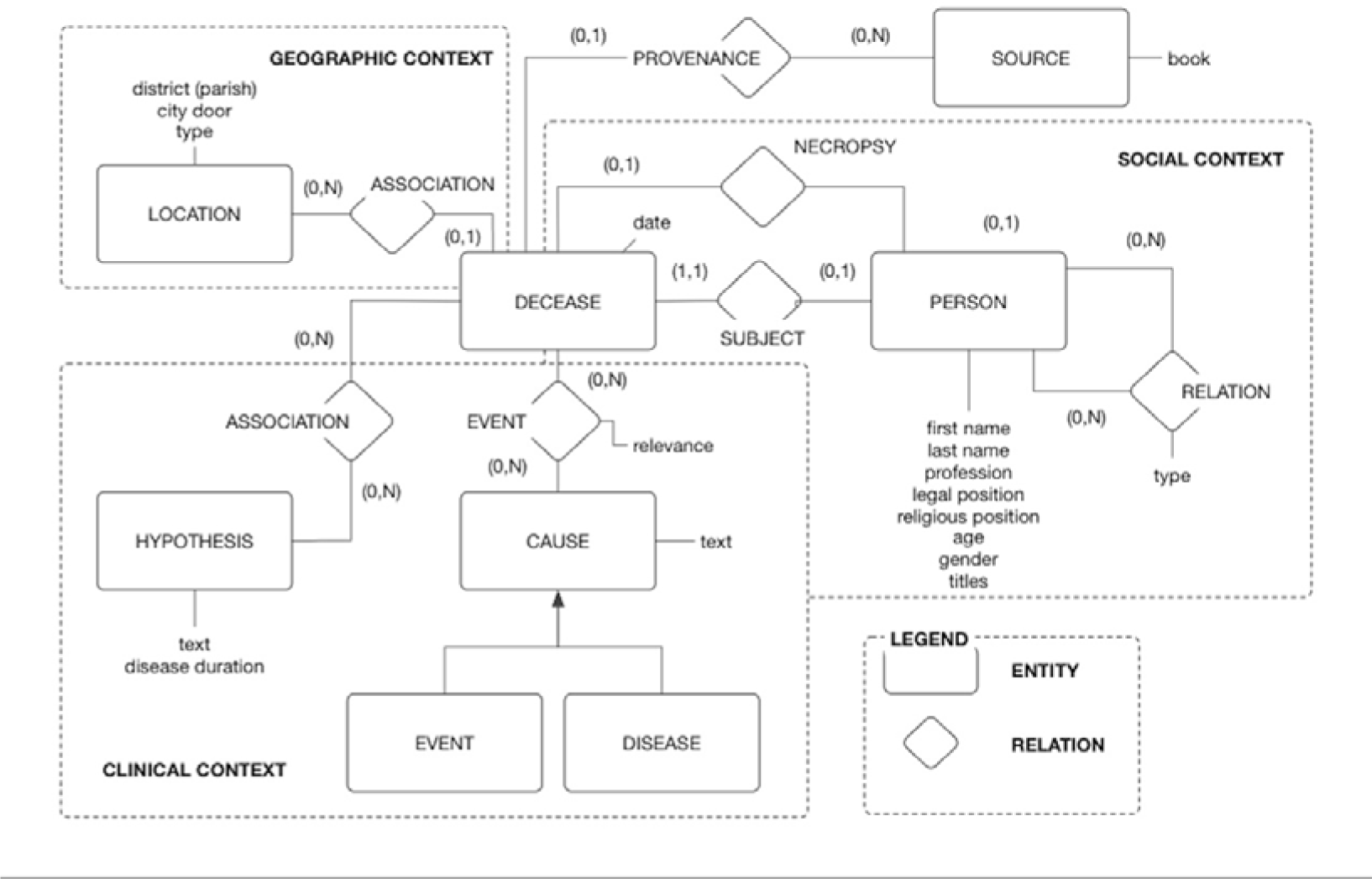
Database structure.

The importance of making historic death register information accessible has been receipted by studies about other death registers projects such as the project based on Bills of Mortality of London, whose data collected from 1664 to 1849 were made accessible to any user as an electronic sheet (UK Data Service, 2020).

An overview of the conceptual structure of the database is shown in Figure 3, where data are described in terms of entities, attributes, relations among entities, and their cardinality constraints. The main entity is the DECEASE, which represents the event of a death as recorded in the MiSfoRe corpus. The clinical context regards the cause of the death and all the concerning information (for example if it was caused by a natural or by a violent cause such as homicide, suicide, or incident). The causes of death were grouped, for analysis purposes, after an accurate interpretation of the terms used in the registers and a careful study of their meaning based on the consultation of medical treatises of the 14^th^ to 16^th^ centuries.

Any recorded death was associated with a date and has a PROVENANCE, that is the reference to the specific book in the corpus where the record has been retrieved. The event can be also associated with the geographical context with the parish and *Sestiere* that are reported in the book and it can also be associated with a description of the socio-demographical condition of the area and, for the last entries of the MiSfoRe, it could be possible to retrieve the type of the houses and some images about the zone. The social context is about the socio-demographic condition of the subject and his or her relatives, it also comprises the relation between subjects (for example a family relationship).

The first entry of the data concerned 1480 (76/I) was made, one of the rare periods of peace and without epidemics. Table 2 contains the variables in the 1480 and their codification. Variables codified by characters were not stored as numeric for the sake of the interrogation of the data, which will be done not only by statisticians but also by historians, anthropologists, doctors, epidemiologists, etc.; the query, for example, will take place by starting to type the name of the parish and through automatic suggestions. For example, it is useful because of the high number of parishes in each year of the registry, as presented for 1480 further on. Missing information was codified as an empty cell to avoid possible problems and misinterpretation in the statistical and epidemiological analyses that can be derived from a numerical codification, for example, 999.

**Table 2.** Variables in the 1480 database and their codification.

| Variables | Codification | Definition (if not defined before) | Levels (if categorical) |
| --- | --- | --- | --- |
| n. | Numeric integer | Progressive number used by identification of subjects |  |
| Register | Character | The identification of the register of the records | 76/I |
| Door | Character |  | Comasina, Nuova, Orientale, Romana, Ticinese, Vercellina |
| Parish | Character |  |  |
| Death Date | Date (yyyy-mm-dd) | The date of death |  |
| Subject | Character | The name, surname or nickname of the deceased person |  |
| Name | Character | The name of the subject (extrapolated from the variable "Subject") |  |
| Surname | Character | The surname of the subject (extrapolated from the variable "Subject") |  |
| Age | Character |  |  |
| Sex | Character |  | F, M |
| Profession of the subject | Character |  | e.g.: canevario, giurisperito |
| Profession of the relative | Character |  |  |
| Legal status |  |  | e.g.: religious, widowed, servant |
| Type of religious | Character | For legal status=religious | e.g.: canon, monk, nun, presbyter |
| Honorary titles of the subject | Character |  | e.g.: domina, dominus, maestro |
| Honorary titles of the relatives | Character |  | e.g.: maestro |
| Name of the relative | Character |  | e.g.: Lorenzo, Ambrogio, Gabriele |
| Degree of kinship | Character |  | e.g.: father, husband |
| Relative state | Character |  | e.g.: dead |
| Type of the place of death | Character |  | e.g.: hospital, castle, monastery |
| Name of the place of death | Character |  | e.g.: *Monastero Nuovo, ospedale di S.Caterina, monastero di S.Francesco* |
| Age_years | Numeric integer | Age of the subjects expressed in years (extrapolated from Age) |  |
| Age_months | Numeric integer | Age of the subjects expressed in months for children and infant, were appropriated (extrapolated from Age) |  |
| Age_days | Numeric integer | Age of the subjects expressed in days for children and infant, were appropriated (extrapolated from Age) |  |
| Tot_age_days | Numeric integer | Calculating from Age_months, Age_years, Age_days |  |
| Cause of death | Character |  |  |
| Physician name (*Necroscopi*) | Character |  | e.g.: *Catelano (Iuditio Catelani), Maestro Ambrogio Pasquali, Maestro Matteo da Busto* |
| Violent death | Indicator variable |  | 0= nonviolent death 1=violent death (suicide, homicide or injuries) |

The database, for now, contains only information about 1480. For now, the database is not available for consultation because the information is still being completed, also as regards external sources.

### The 1480 register: epidemiological analysis

1480 is one of the rare periods of peace and without epidemics and included 1813 subjects. This section describes the age and sex distribution of the deaths observed in 1480, to contribute to depicting an epidemiological and demographic profile of Milan in the last part of the 15^th^ century. For this purpose the number of deaths that occurred in each parish was assessed, to compare the number of cases observed in the different areas of the city. The deaths in the extreme ages (childhood or old age) in the absence of a specific attribution to a disease, or related to childbirth, or deaths from starvation had been also investigated as well as the deaths from unnatural or violent causes and the deaths that occurred in the hospitals of the city.^7^

To render the data readable by software such as R (R Core Team 2021) and to conduct statistical and territorial analysis, the following steps were performed on the first input of the data^8^:

1. The repetition of the same records (full individual information corresponding to a row of the database) has been removed.
2. The presence of special characters, such as typos, asterisks, or non-Latin letters, in the header and the cell was checked and removed. The recordings of missing data, when a variable is missing for a particular registered death were codified as an empty cell, as NA. If the information about the missing data could be deduced from other fields, a correction has been made (i.e.: a subject had a NA for Sex; the name of this subject was *Giovannina*, so the Sex could be coded as female).
3. A spelling or typing error correction has been performed when it could be deduced from other fields (i.e.: A*. Maria al Circolo*, in the field about the parish, has been corrected as *S. Maria al Circolo* because the first was an exception with respect to the rest). Those cases where the inconsistencies or errors could not be attributed to a typing were solved after an expert check the manuscript.

The mortality registrations in 1480 referred to many parish churches which no longer exist; the location of these churches has been identified as precisely as possible. In particular thanks to some material found online (Di Bello 2017) it was possible to localize those churches, they then were positioned with QGIS on the map of Milan. Several parishes were divided into two parts, one inside (*intus*) and one outside the second (medieval) ring of walls (*foris)* (for example, *San Lorenzo foris* and *San Lorenzo intus*). Each subject had a record about the parish, with the specification about the part of the parish the record is referred to, if it was not specified, *foris* or *intus*, the record was considered as referred to the *intus* part of the parish in the case the churches was in the area defined by the Medieval walls and as *foris* otherwise.

Most of the parishes were concentrated in the central part of the city and were very near each other, so the representation of the distribution of death in them would potentially lead to a result that is difficult to interpret. On the other hand, the *Sestieri* were relatively large, and the territory of each of them starts from the center to the extreme periphery of the city, thus including a heterogeneous population. On the contrary, it was tentatively conceivable that any *Contrada* had a relatively homogeneous population from a socio-economic point of view. Thus, for analysis purposes, the deaths that occurred in the parishes that persisted in the territory of each *Contrada* were grouped. Moreover, the number of deaths that occurred in the *Contrade*, inside the old walls, was compared to that observed in the parishes placed between them and the third circle of walls which was under construction at the time of the Sforzas. The territorial density of deaths occurred in the area of competence of each *Contrada* and the entire area between the medieval ring and the new wall ring was expressed as (number of events/area of the Contrada expressed in square meters) * 1000. The analyzes were performed using Qgis (vers. 3.16.2) (Qgis Development Team 2021) utilizing OpenStreetMap (© OpenStreetMap contributors) as a base map.

### The deaths registered in Milan in 1480

Mortality may be an important indicator of the size and socioeconomic status of the population living in each area of the city. After excluding some double registrations, the 1480 register included, starting from January 23^rd^, a total of 1813 death reports. It was impossible to find the recordings of the first 22 days, which would have been accessible in the 19^th^ century to Ferrario (Ferrario 1840), who reported for January 1480 a total of 170 deaths, and a total of 1938 deaths in the entire year. Table 3 compares the monthly deaths in 1480 of our data to those reported by Ferrario (Ferrario 1840). Only minimal differences were evidenced, apart from the deaths that occurred in January. No significant differences have been observed in mortality in the different seasons.

**Table 3.** Deaths monthly registered in 1480 as reported by Ferrario compared to those found in the present study.

| Month | Jan | Feb | Mar | Apr | May | Jun | Jul | Aug | Sep | Oct | Nov | Dec | Tot |
| --- | --- | --- | --- | --- | --- | --- | --- | --- | --- | --- | --- | --- | --- |
| Ferrario | 170 | 190 | 144 | 194 | 162 | 106 | 191 | 192 | 130 | 159 | 164 | 146 | 1948 |
| Present study | 50 | 178 | 150 | 194 | 161 | 107 | 190 | 192 | 131 | 159 | 153 | 148 | 1813 |

Table 4 presents the distribution of age at death in years by gender of the 1,811 subjects for whom age was reported. Females had a slightly older age at death (26,78 years in mean and 18 years in median) than males (25,55 years in mean and 14 years in median). Other measures (minimum, maximum, first, and third quartile) are very similar for females and males, so the dispersion of the distribution of age between the two genders is very similar.

**Table 4.**
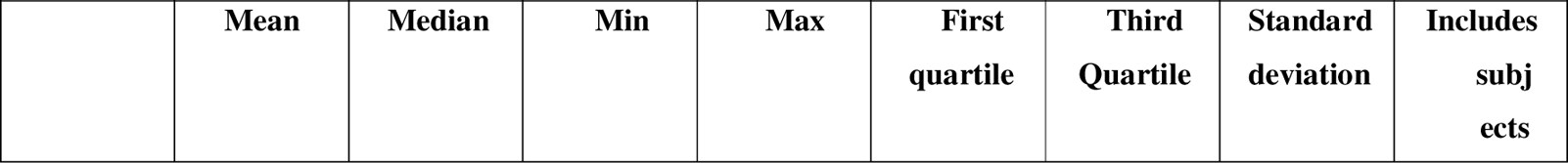
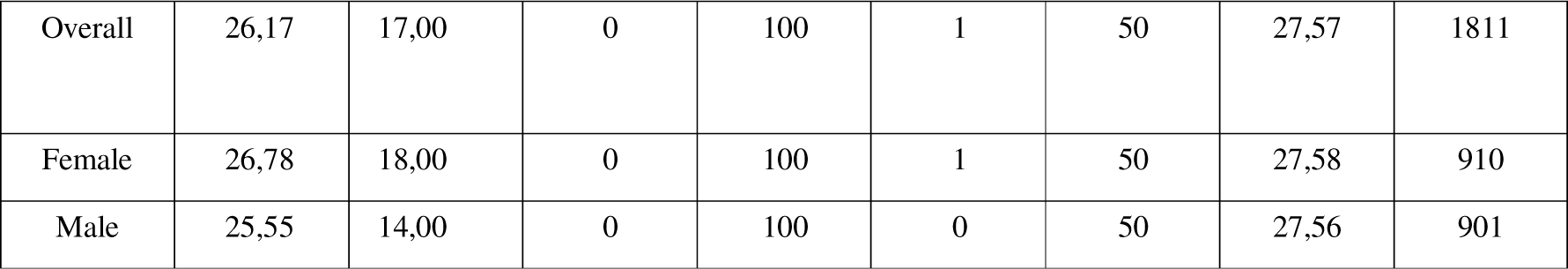
Age (years) at death and distribution by gender.

|  | Mean | Median | Min | Max | First quartile | Third Quartile | Standard deviation | Includes subjects |
| --- | --- | --- | --- | --- | --- | --- | --- | --- |
| Overall | 26,17 | 17,00 | 0 | 100 | 1 | 50 | 27,57 | 1811 |
| Female | 26,78 | 18,00 | 0 | 100 | 1 | 50 | 27,58 | 910 |
| Male | 25,55 | 14,00 | 0 | 100 | 0 | 50 | 27,56 | 901 |

However, the mortality by gender was not significantly different in the different age classes (figure 4). Women have a longer life expectancy than man also in modern times and also in conditions of famine, slavery, and epidemics (Zarulli et al. 2018). About 41% of the total deaths in males and 39% of those in females occurred in children aged 0-5 years.

**Fig. 4.**
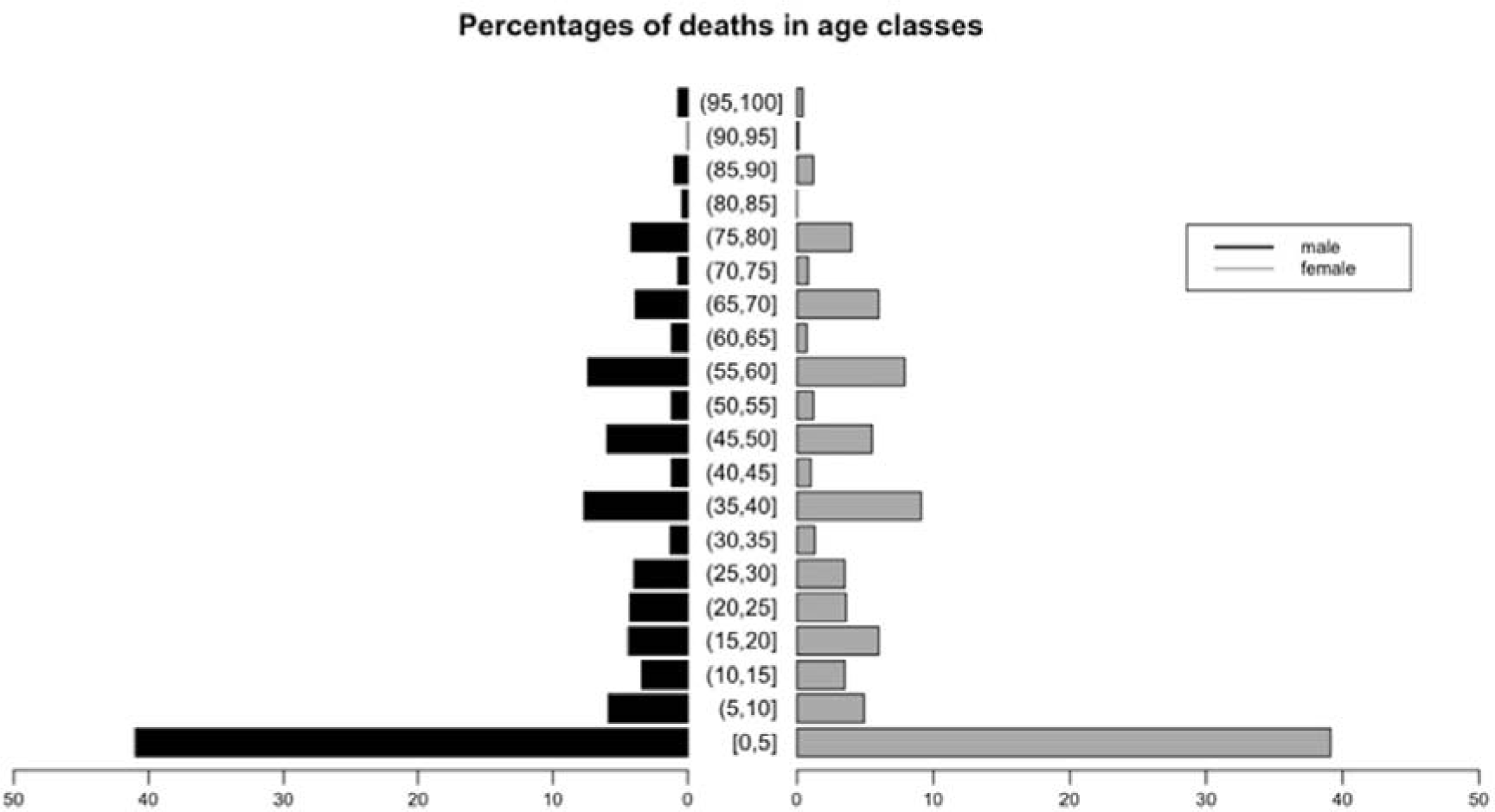
Comparison of percentages of deaths in the different age classes in males and females.

Children’s mortality (under 10 years of age) is in connection with the life expectancy in the society: it has been estimated that if the latter is less than 30 years, then 40% of mortality will occur in children; if the life expectancy is 50 years, the children mortality will be 20% and if it is above 60 years, children mortality will be 10% (Woods 2006). In the Milan of 1480, according to this historical source, the median age at death for adults was 40 years as shown below.

In the registry, only for perinatal subjects, there are also an indication about the months and the days of the age at death, but we rounded them in the nearest years. In general, the age at death, in the registration was approximated and this was especially the case for older people in which there were a lot of values terminated with a zero (Zanetti 1976).

A round bracket indicated an open interval and a square bracket indicated a close interval, so, for example, the notation [0, 5] indicates that the interval includes subjects who have at least zero years and have a maximum of five years (so included the subject with zero years and five years). The (5, 10] includes subjects that have more than five years and a maximum of ten years (so included the subject with ten years and excluded the subjects with five years)

Interestingly, among the 1,046 subjects remaining in the analysis after excluding children younger than 8 years old, no further differences in the age at death between males and females were observed (Table 5). In particular, for females, the mean was 44,32 years and the median was 40 years and for males, the mean was 44, 60 years and the median was 40 years. Once again the dispersion measures testify that the dispersion of the distribution of age of the two genders is very similar.

**Table 5.** Distribution of age (years) at the death after excluding infant mortality.

|  | **Mean**<br>**(years)** | **Median**<br>**(years)** | **Min**<br>**(years)** | **Max**<br>**(years)** | **First**<br>**quartile**<br>**(years)** | **Third**<br>**Quartile**<br>**(years)** | **Standard**<br>**deviation**<br>**(years)** | **Included**<br>**subjects** |
| --- | --- | --- | --- | --- | --- | --- | --- | --- |
| All subjects | 44,45 | 40,00 | 8,00 | 100,00 | 24,00 | 60,00 | 22,84 | 1046 |
| Female | 44,32 | 40,00 | 8,00 | 100,00 | 24,00 | 60,00 | 22,85 | 540 |
| Male | 44,60 | 40,00 | 8,00 | 100,00 | 25,00 | 60,00 | 22,84 | 506 |

Male’s over mortality could be explained by genetic causes (biological hypothesis): having xy chromosomes make the males more susceptible to recessive diseases on the x chromosome because the second x chromosome is not present as a counterpart that makes recessive traits non-expressible. There are some environmental factors that affect the health of the baby both in the uterus and after birth: parental stress, medical conditions during conception, occupation, and exposure to environmental risks; these conditions particularly affected males. An analysis that involved 50.000 twins in sub-Saharian African countries tested these two hypotheses on a male over mortality and gave results that affirm their role. Twins were chosen to conduct the analysis because they are exposed to the same conditions (Pongou, Kuate Defo, and Tsala Dimbuene 2017).

#### The distribution of deaths in the different parts of the city

The parishes cited in the register were 104, a large number for a relatively small territory, mainly concentrated especially in the central part of the city. The number of deaths observed in each parish was very variable, probably reflecting the different size, populousness and social conditions of each of them.

It was not possible to determine, also in approximation, the distribution of the population of Milan in the different parts of the city because ancient maps, to our knowledge, in the medieval and Renaissance period were few and extremely idealized.^9^

So it is assumed an equal distribution of the population in different parts of the city, also because in our knowledge there isn’t a document about this.

Figure 5 shows the location on the map of Milan of each church and corresponding parish in which a larger than a usual number of deaths occurred (defined as deaths higher than 1% of the total mortality observed).

**Fig. 5.**
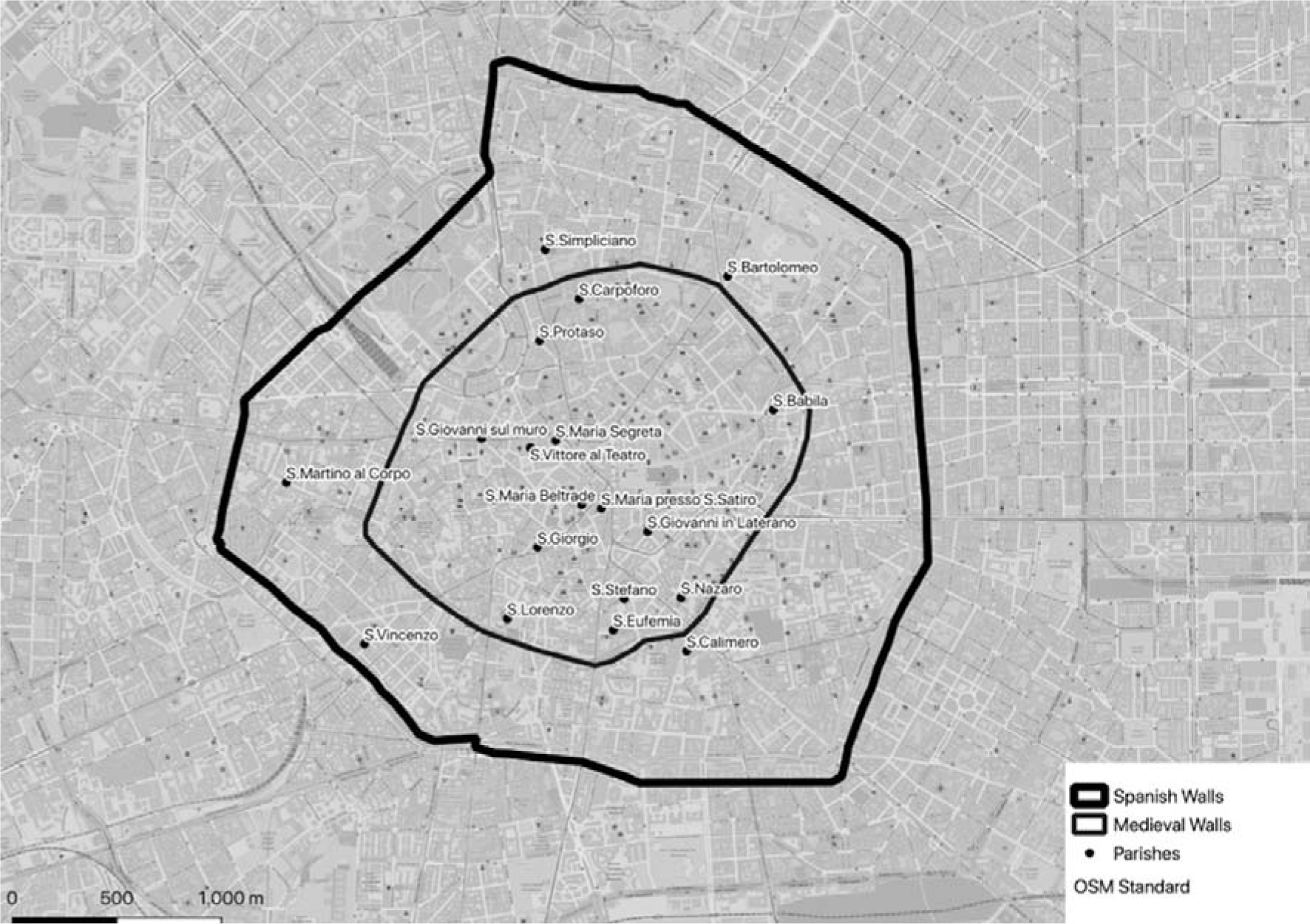
Distribution on the Milan map of the parishes with more than 1% of the total number of deaths.

Table 6 shows the percentage of the total mortality that occurred in each parish. The highest mortality was seen in the parishes of *S. Lorenzo foris, S. Protaso foris, S. Simpliciano foris, S. Nazaro intus* and *S. Calimero foris*.

**Table 6.** Percent distribution of deaths higher than 1% of total mortality by parish.

| Parish | % (N=1813) |
| --- | --- |
| S.Lorenzo foris | 10,4 |
| S.Protaso foris | 5,6 |
| S.Simpliciano | 5,4 |
| S.Nazaro | 5,2 |
| S.Calimero | 5,1 |
| S.Bartolomeo | 4,3 |
| S.Stefano foris | 3,9 |
| S.Stefano intus | 3,5 |
| S.Babila intus | 3,3 |
| S.Babila foris | 3,1 |
| S.Martino al Corpo | 2,9 |
| S.Protaso intus | 2,9 |
| S.Carpoforo intus | 2,2 |
| S.Eufemia intus | 2,2 |
| S.Maria Beltrade intus | 1,8 |
| S.Vincenzo foris | 1,7 |
| S.Giorgio | 1,5 |
| S.Lorenzo intus | 1,5 |
| S.Eufemia foris | 1,4 |
| S.Vittore al Teatro | 1,3 |
| S.Giovanni in Laterano | 1,2 |
| S.Giovanni sul muro | 1,2 |
| S.Maria presso S.Satiro | 1,2 |
| S.Maria Segreta | 1,1 |

Figure 6 illustrates the distribution of the deceased people in the Contrade taking into consideration the area of each Contrada and in the area between the Spanish walls and the medieval walls. The number and the percentage of the deceased people among the Contrade and outside the *Contrade* are in Table 7. The high density of events in *Contrade* 21, 16, 12, 11, and 3 could reflect their larger population compared to the others. In the 16^th^ and 17^th^ centuries, the poorest segments of the population were distributed throughout the city, with greater density in the more peripheral parts and some areas of the center (D’Amico 1995). There is no information available in this regard on the situation in the 15^th^ century, but it can be assumed outside the medieval walls resided the majority of the poorest sections of the population.

**Fig. 6.**
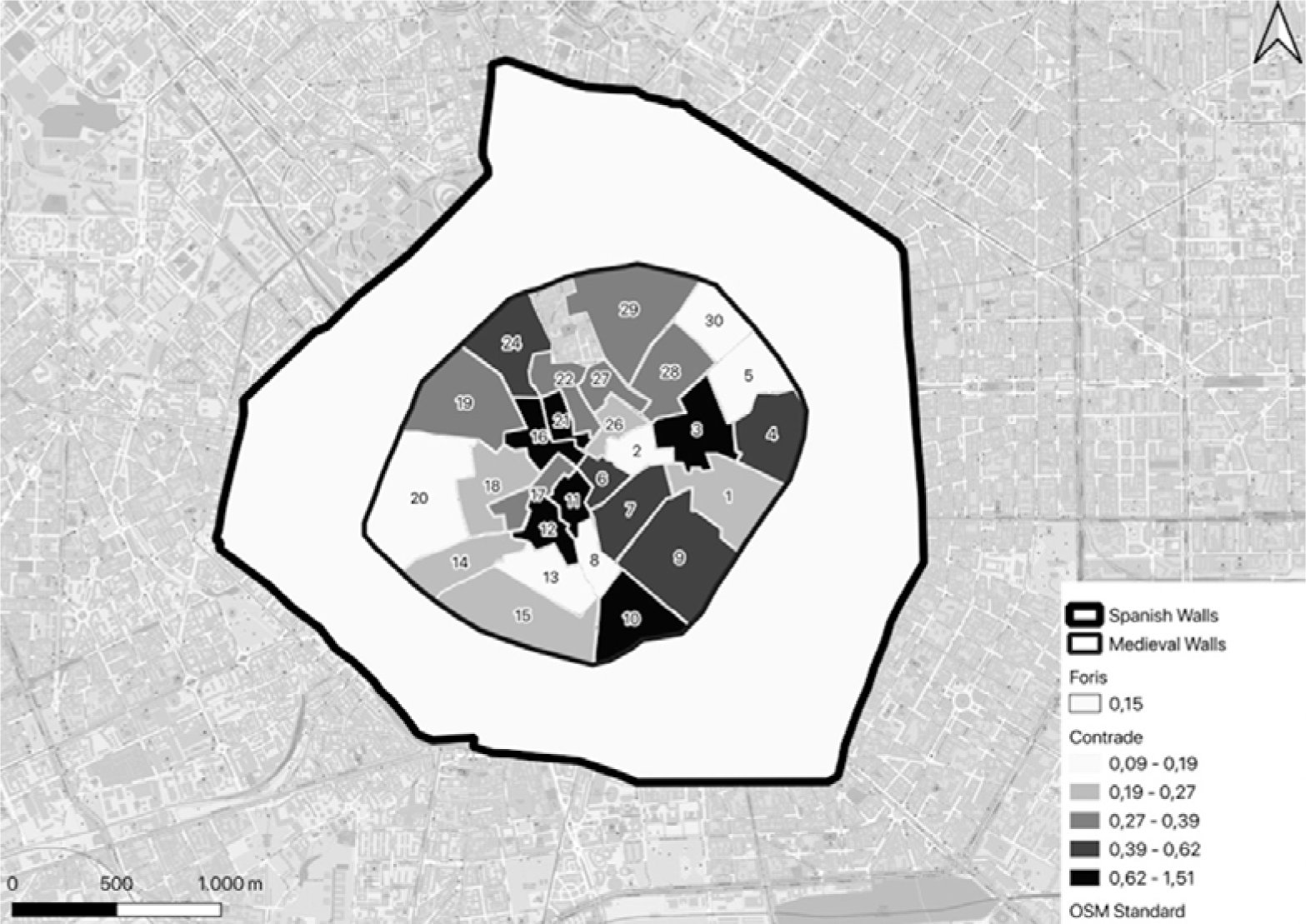
Distribution of the deceased people in the Contrade taking into consideration the area of each Contrada and in the area between the Spanish walls and the medieval walls.

**Table 7.** The number of deaths and the percentage of the total deceased persons in each *Contrada* and the area outside the *Contrade*. The denominator was the subjects in the registry.

| ID (Name) | N= 1813 (%) |
| --- | --- |
| *INTUS* | 54,8 |
| *9 (Contrada del Brolo)* | 6,5 |
| *10 (Contrada delle Capre)* | 5,6 |
| *3 (Contrada dell'Agnello)* | 3,5 |
| *4 (Contrada della Cerva)* | 3,3 |
| *24 (Contrada del Campo)* | 2,9 |
| *29 (Contrada degli Andegari)* | 2,9 |
| *19 (Contrada della Porta)* | 2,8 |
| *15 (Contrada della Vetra)* | 2,7 |
| *7 (Nobile Contrada della Cicogna)* | 2,6 |
| *11 (Contrada della Lupa)* | 2,6 |
| *21 (Nobile Contrada del Cordusio)* | 2,4 |
| 16 (*Contrada della Piscina*) | 2,1 |
| 28 (*Contrada della Mazza*) | 2 |
| 12 (*Nobile Contrada di Sant'Ambrogio*) | 1,8 |
| 1 (*Contrada del Verzaro*) | 1,5 |
| 18 (*Contrada dei Morigi*) | 1,2 |
| 20 (*Contrada del Nirone*) | 1,2 |
| 14 (*Contrada del Torchio*) | 1 |
| 22 (*Contrada del Rovello*) | 1 |
| 5 (*Contrada di Bagutta*) | 0,8 |
| 6 (*Contrada del Falcone*) | 0,8 |
| 17 (*Nobile Contrada della Rosa*) | 0,8 |
| 27 (*Nobile Contrada dei Bossi*) | 0,8 |
| 13 (*Contrada delle Cornacchie*) | 0,4 |
| 26 (*Contrada Capitana*) | 0,4 |
| 30 (*Contrada della Spiga*) | 0,4 |
| 2 (*Nobile Contrada delle Farine*) | 0,3 |
| 8 (*Contrada del Fieno*) | 0,2 |
| **FORIS** | 45,2 |

So it is possible that the high mortality in parishes outside the medieval walls reflects this fact. For parishes inside the medieval walls, high mortality may be due to a high density of population. Overall, there were 994 (54,8%) deaths occurred in the areas inside the medieval walls) and 819 (45,2%) outside (table 7).

#### Distribution of the causes of death

Table 8 shows the percentage of death by cause. The deaths attributed to a defined disease are grouped.

**Table 8.** Deaths occurred in 1480 grouped by identified causes.

| **Causes of death** | **% (N=1813)** |
| --- | --- |
| Perinatal mortality | 19,2 |
| Children deaths, no causes reported | 16,2 |
| Childbirth or related causes | 1,7 |
| Old age | 1,2 |
| Unknown/ undetermined/ unreadable | 1,2 |
| Starvation | 0,4 |
| Other causes of death | 58,6 |
| **Not natural/Violent causes of death** |  |
| Accident | 0,8 |
| Homicide | 0,7 |
| Suicide | 0,2 |
| Drowning | 0,1 |

As will be discussed in more detail below, health authorities paid limited attention in the register to the causes of infant death. Thus, perinatal mortality, the single most frequently-cited ’cause’ of death was reported without any further specification as so as the deaths of the large majority of children.

#### Infant deaths

The total number of deaths registered for children 0 to 7 years old was 765, of which 395 (51,6%) were observed in males and 370 (48,4%) in females. Table 9 shows the distribution of age at death in infants.

**Table 9.**
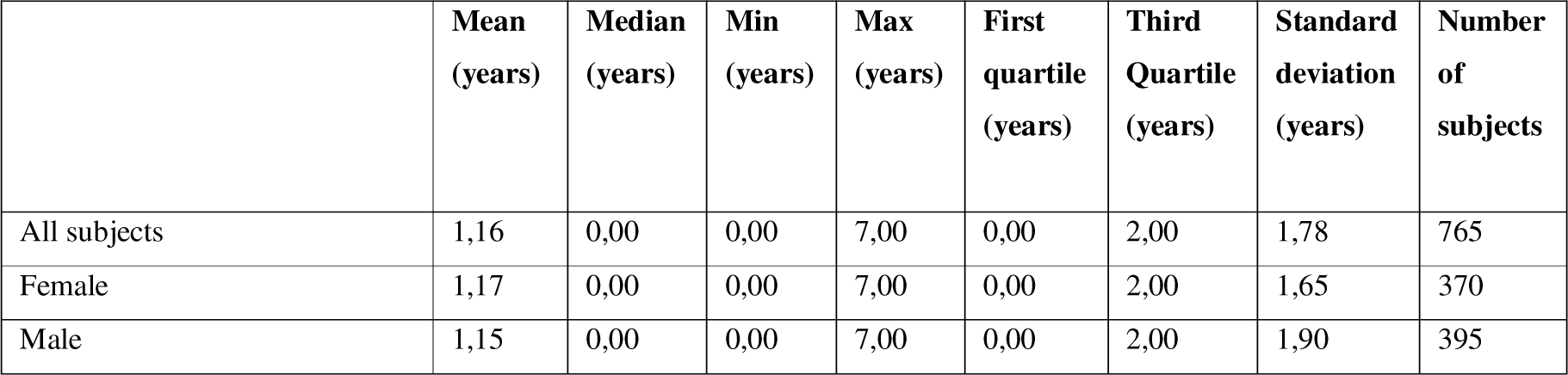
Distribution of age (years) at death for children aged 0 to 7

About 50% of the infant deaths occurred within the first year of life, and 75% before the end of the second. The median and the mean ages at death for females and males are very similar. These data on infant mortality in Milan in 1480 are to our knowledge the oldest available today among those taken from death registers. It is known that the deaths observed in the first 27 days of life should be mainly attributed to ‘endogenous factors’ such as inborn diseases, congenital abnormality, birth trauma, or pre-term birth, while deaths occurring from the 28^th^ day to 1 year of age are more likely to ‘exogenous causes’, such as infectious disease, malnutrition and poor living conditions (Bourgeois-Pichat 1951; Lewis and Gowland 2007). A higher risk of mortality of males during childhood occurs also in modern times. In Italy in 2016 the mortality of males was 3 for one thousand live births males and females it was 2,6 (ISTAT 2019).

Table 10 shows the age at death in infants, i.e. in children who died younger than 28 days.

**Table 10.** Age at death in infants (subjects with less than 28 days at death)

|  | **Mean**<br>**(days)** | **Median**<br>**(days)** | **Min**<br>**(days)** | **Max**<br>**(days)** | **First**<br>**quartile**<br>**(days)** | **Third**<br>**Quartile**<br>**(days)** | **Standard**<br>**deviation**<br>**(days)** | **Number**<br>**of**<br>**subjects** |
| --- | --- | --- | --- | --- | --- | --- | --- | --- |
| All subjects | 5.80 | 6 | 0 | 21 | 1 | 8 | 4.86 | 233 |
| Female | 6.59 | 7.5 | 1 | 21 | 1 | 8 | 5.31 | 94 |
| Male | 5.27 | 4 | 0 | 20 | 1 | 8 | 4.47 | 139 |

These observations are similar to those reported in recent times in disadvantaged populations in low-income countries; in a study conducted in Ghana about data between January 2003 and December 2009, the 64,8% (of 424 neonatal deaths) occurred in the first week of life (Welaga 2013). An excess of mortality in male infants has been well described both in contemporary and historic populations (Stinson 1985; Drevenstedt et al. 2008) In particular, the relative fragility of male infants was confirmed by an analysis of data from fifteen developed countries regarding the period between the mid-18^th^ century and 1970, where the excess of male mortality increased in association with an overall decrease in infant mortality. The male disadvantage could be related to the decline of the contribution of infectious disease to infant mortality; in this case, infant mortality was increasingly the result of compromised perinatal conditions, conditions which affected boys to a greater extent. The decrease in this male disadvantage after 1970 was due to the decline in the contribution of perinatal conditions to infant mortality (Drevenstedt et al. 2008).

The distribution of deaths under the age of eight, stratified by age and by parish is reported in Table 11.

**Table 11.**
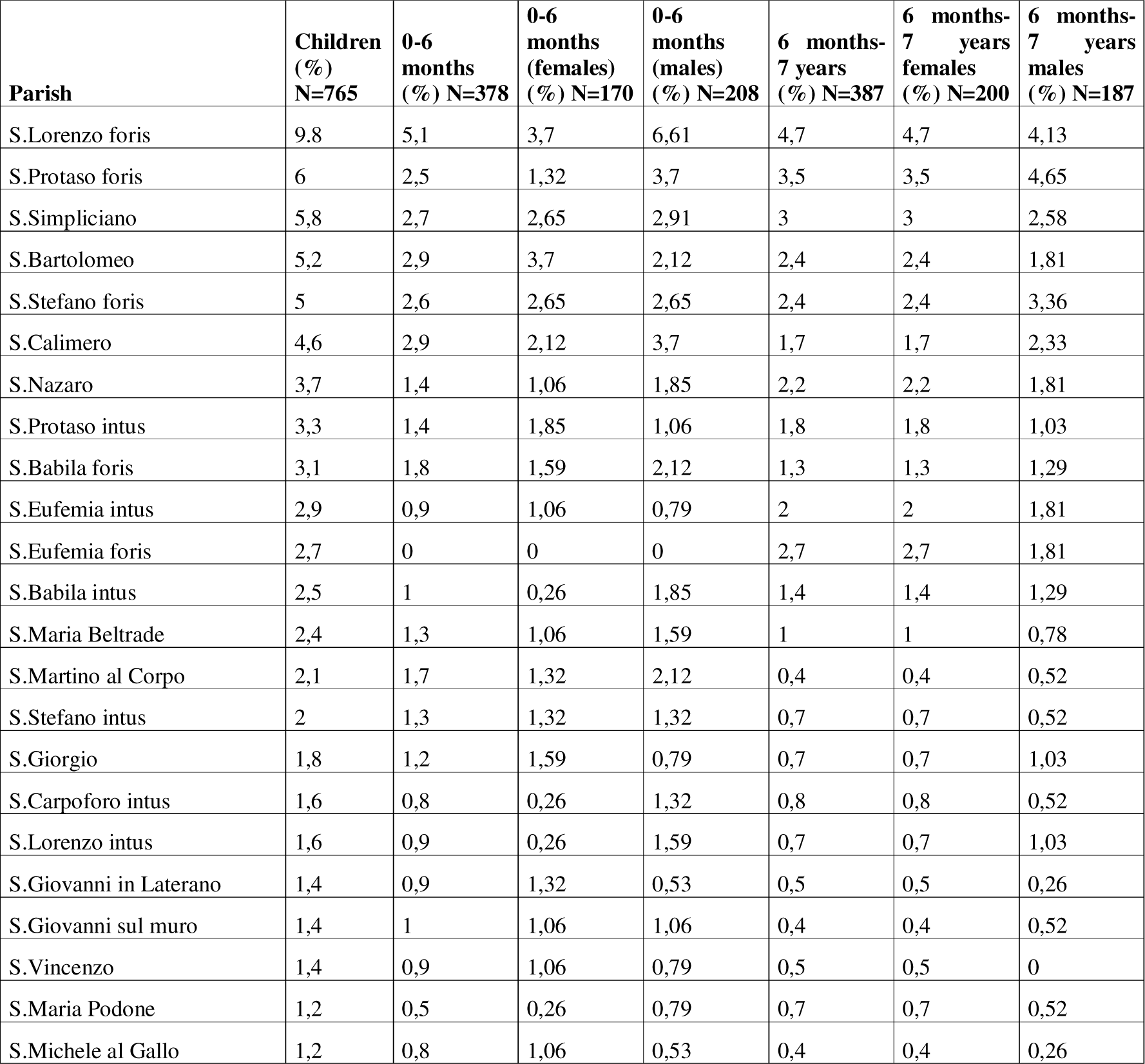
Distribution of subjects from 0 to 7 years old in parishes. Only the parishes reaching with at least 1% of the distribution total of the death occurred in of children from 0 to eight years old were considered. Percentages are expressed by columns.

The parishes with the highest perinatal and infant mortality were *S. Lorenzo foris* (9.8% of children mortality)*, S. Protaso foris* (6% of children mortality), and *S. Simpliciano* (5.8% of children mortality). All of them were located outside the ring of medieval walls, in the peripheral part of the city (Fig. 7), where it can be presumed a high density of disadvantaged people and wage earner workers and this fact can be seen also in the children’s mortality; mortality, in general, is an indicator of the socio-economic level and in particular the rate of infant mortality (Haines 1995).

**Fig. 7.**
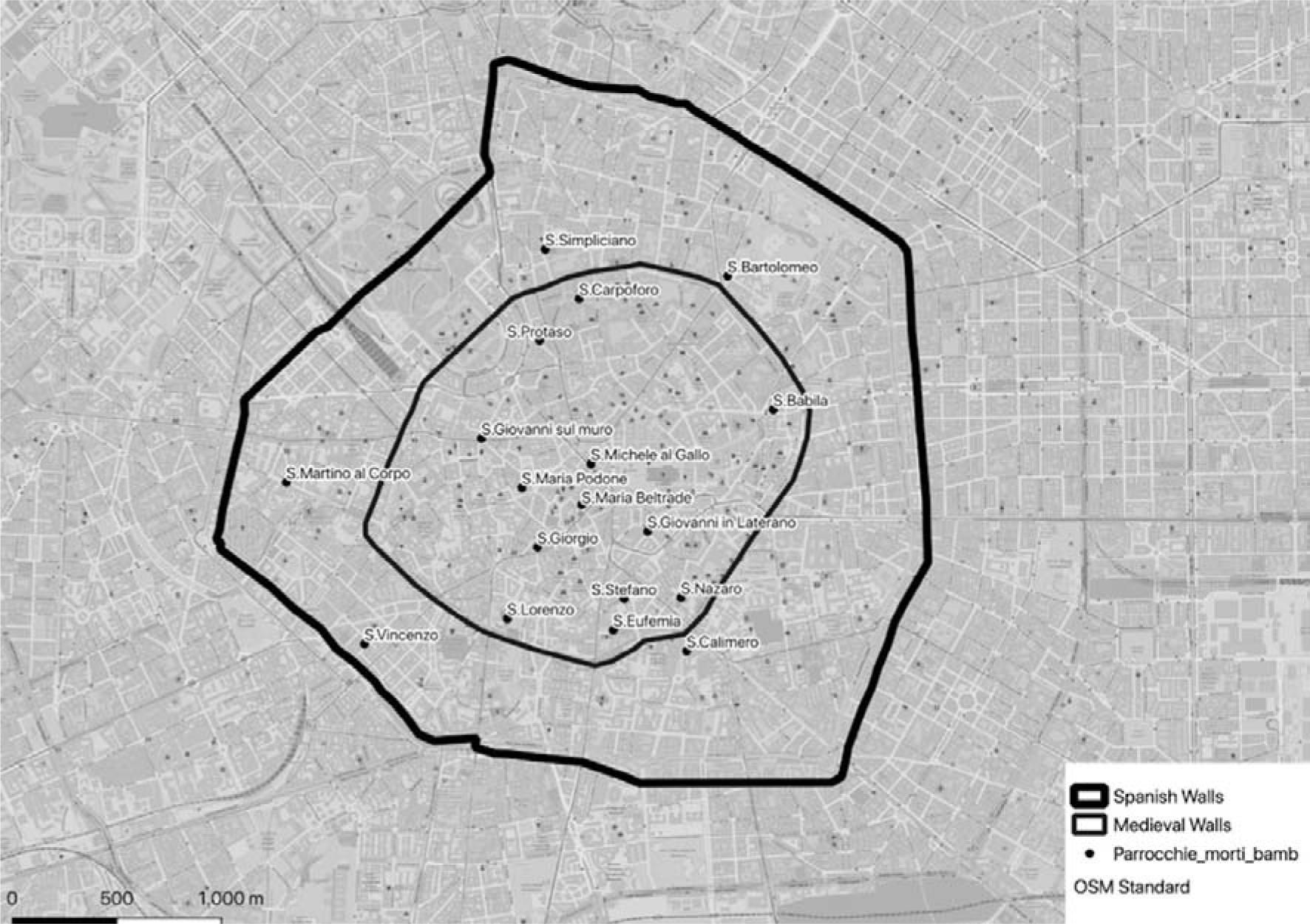
Distribution on the Milan map of the parishes with an infant and perinatal mortality more than 1% of the total number of deaths under eight years of age.

In London during from 1580 to 1650 a higher male infant mortality was observed in poorer parishes, but not in the wealthier ones (Finlay 1981). Similarly, in the parish of Christ Church Spitalfield in London there was significant excess male infant mortality in the 1750–59 cohort, but not in later ones (Humphrey, Bello, and Rousham 2012).

Figure 8 shows the distribution of the deaths under eight years of age in the different parts of the city.

**Fig. 8.**
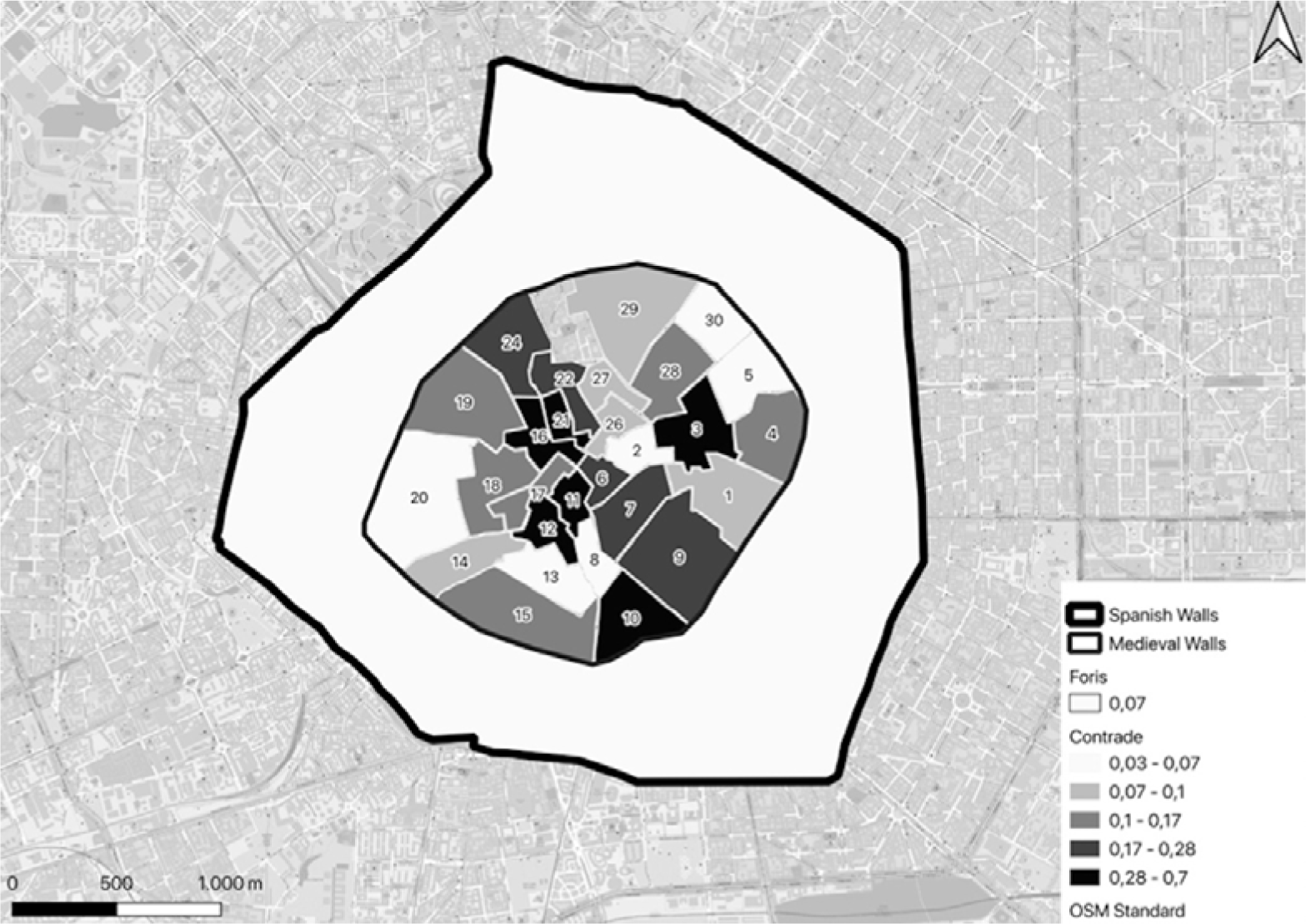
Distribution of the deceased from 0 to 7 years old among the *Contrade* and in the area outside the *Contrade*. The value number of deceased in each part of the city considered was divided by the surface in square meters of each of them and then multiplied by 1000.

The number and the percentage of children’s deaths in each *Contrada* and in *foris* area are shown in Table 12. The distribution reflects that of the total deaths registered. Also, this data suggests that a high number of deaths, regardless of age at death, account not only for the populousness of the area concerned but also for the level of indigence of the inhabitants.

**Table 12.** Number of subjects from 0 to 7 years old and percentage of total observed deaths in each Contrade and the area outside the Contrade (foris).

|  | **TOT=765** |
| --- | --- |
| **ID CONTRADA (Name)** |  |
| 9 (*Contrada del Brolo*) | 5,1 |
| 10 (*Contrada delle Capre*) | 4,8 |
| 3 (*Contrada dell'Agnello*) | 4,4 |
| 11 (*Contrada della Lupa*) | 3,3 |
| 24 (*Contrada del Campo*) | 3,3 |
| 15 (*Contrada della Vetra*) | 2,7 |
| 19 (*Contrada della Porta*) | 2,6 |
| 4 (*Contrada della Cerva*) | 2,5 |
| 29 (*Contrada degli Andegari*) | 2,5 |
| 7 (*Nobile Contrada della Cicogna*) | 2,2 |
| 12 (*Nobile Contrada di Sant'Ambrogio*) | 2,2 |
| 16 (*Contrada della Piscina*) | 2,1 |
| 21 (*Nobile Contrada del Cordusio*) | 2 |
| 28 (*Contrada della Mazza*) | 2 |
| 1 (*Contrada del Verzaro*) | 1,4 |
| 18 (*Contrada dei Morigi*) | 1,3 |
| 20 (*Contrada del Nirone*) | 1,3 |
| 22 (*Contrada del Rovello*) | 1,3 |
| 6 (*Contrada del Falcone*) | 1,2 |
| 14 (*Contrada del Torchio*) | 1 |
| 17 (*Nobile Contrada della Rosa*) | 0,7 |
| 5 (*Contrada di Bagutta*) | 0,5 |
| 13 (*Contrada delle Cornacchie*) | 0,5 |
| 27 (*Nobile Contrada dei Bossi*) | 0,5 |
| 26 (*Contrada Capitana*) | 0,4 |
| 30 (*Contrada della Spiga*) | 0,4 |
| 2 (*Nobile Contrada delle Farine*) | 0,3 |
| 8 (*Contrada del Fieno*) | 0,1 |
| **FORIS** | **47,3** |
| **Total number of children** | **765** |

#### Mortality in foundlings

The register reports a total of thirty deaths at the San Celso hospital, the foundlings’ hospice^10^. These deaths occurred in children aged between 2 and 9 years (median 3 years) whose foundling condition is confirmed by the surname *da Milano* common to all of them. Even in the absence of data regarding the number of guests, allowing us to estimate the mortality, it is likely that mortality among foundlings was very high (Cernik 2016; D’Andrea and Terpstra 2017).

Unlike what was observed for deaths in children occurring at their home, for these foundlings, the cause of death was reported in all but three cases, for whom the name of the author of the certification is also missing. On the other hand, 25 certifications were given by the same doctor, Maestro Giovanni Casetti, probably the doctor in charge of hospital care. The most frequent causes of death reported are ethical fever with 11 cases without other association and other 5 associated with other diseases. Ethical fever was a chronic fever ‘generated by bones’^11^ an often associated with phthisis or with long-lasting inflammatory illnesses. The second most frequent cause of death was in 10 subjects a diarrheal diseases with or without fever. The distribution during the year of such cases does not suggest the occurrence of an epidemic cluster. Four deaths were attributed to generalized edema, which in this case might be attributed to protein deficiency due to malnutrition.

#### Childbirth mortality

Among all deaths registered in Milan in 1480, 30 concerned women died of childbirth and/ or related causes (1,7% of the total cause of death and 3.3% of causes of death of females). The deaths in women in childbearing age (15-45 years) were 229 (25.2% of the total deaths in women). The mean age of those women at death was 29.83 years, the median was 30 (sd=9,21). Among the women who died from childbirth the median age was 28.5 years (sd= 5.81, min=20 years, max=40 years); 12 of them (40%) resided within and 18 (60%) outside the medieval walls. Considering all women who died during childbearing age (15-45 years), 120 (52,4%) died within and 109 (47,6%) outside the medieval walls.

In *S. Lorenzo foris* six childbirth deaths have occurred, one-fifth of the total, further suggesting the probable populousness and possible poverty of that parish.

#### Deaths of old age and causes of death in elderlies

The register reports 21 deaths of old age (9 males and 9 females, median age 90 years, range 80-100) without reporting causes other than this. The age in these cases has been presumptively attributed, with an approximation to ten in almost all of them. Only one of them deceased in a hospital and five were supposed to have at death 100 years or more. The centenarians whose death from any cause is recorded in the register of 1480 were seven in all, three males and four females. Overall, deaths aged 70 and over were 205 (11,3%).

#### Starvation

Six cases between the ages of six and 60, five of which are female, were reported in the register by Mastro Dionigi from Nuremberg with the formula “found undernourished and consumed”. In a case, it was also specified that the visit had occurred *post-mortem*, as probably happened in all these cases. It is therefore probable that these deaths occurred from starvation in people who had not had access to means of subsistence and had not received assistance before death. Therefore, it can be supposed that starvation was the cause of at least 0,3% of the deaths that occurred in Milan in 1480.

#### Not natural/Violent causes of death

There were 33 non-natural/violent deaths in the dataset, 1,8% of the total, a percentage almost identical to that detected by Dante Zanetti in the MiSfoRe of 1503 (55 cases out of 3156 deaths, 1,74%) (Zanetti 1976). Interestingly, in 2012 the percentage of non-natural deaths on the total recorded deaths in Italy was slightly higher (2,27%) and becomes quite similar if the deaths that occurred in transport accidents are subtracted from the total (ISTAT 2012). A marked difference, however, was seen in the deaths from murder, in the Italy of 2012 were the 0,07% of the total deaths recorded, with about three-quarters female victims. On the contrary, in the Milano of 1480, the homicides represented 0.66% of the total deaths and all the murdered were males of the median age of 27 years (range 17-50). Although it cannot be excluded that violent deaths of women at any age may have been hidden under other apparently natural causes, it seems to be possible to conclude that femicide was not frequent in Milan at the end of the fifteenth century. In the study we are completing in the 1629 register, out of 59 murder cases, only one involved a woman. All of the men murdered in 1480 were killed by sidearms, probably as a result of stabbings or duels with swords^12^.

Due to the exclusion of suicides from burial in consecrated land, some of the deaths from suicide might have been registered as accidental, as in the case of an 18-years-old girl, registered on January 30, who died from a ‘fall from above’. An attempt to justify a suicide with a mental illness is that of *Johannina famula* (servant of) *Magdalene de Grulis,* an 18-years-old girl, registered on June 24^13^. The health officers were not so polite with another 21-year-old girl registered on February 12^14^.

Among the deaths due to accidental causes, two deserve to be mentioned as an example of the accuracy of the registrations in the MiSfoRe. The first is the case of *Vincenzo di* (son of) *Ambrogio Griffi*, 23 years old *qui balando cecidit super tavolam cum capite et facta forti contusione est cerebrum in 7ª* (dancing he fell on a table with his head and got a severe contusion to the brain, and died in the 7^th^ day). The second is the accident that occurred to *Giacomo Pozzobonelli*, a 96 years old man who dead *ex casu ab alto cum fractura cosse sinistre et contusione totius corporis* (for a fall from above with a fracture of the left thigh and bruising of the whole body)^15^.

#### Deaths in hospitals

In addition to the 30 foundlings who died in *San Celso*, another 122 deaths occurred in a total of 11 other hospitals. Seven of them (*San Dionigi*, 9 deaths; *San Vincenzo*, 5; *San Lazzaro* and *Santa Caterina*, 4 each; *San Giacomo* and *San Simpliciano*, 3 each; *Ospedale Nuovo*, 2) probably functioned as parish hospitals or hospices, with the only exception of *San Lazzaro*, dedicated to the treatment of wounds and skin diseases. Of the other four, the two largest, the *Ospedale Maggiore* and the *Brolo* hospital recorded 32 and 31 deaths respectively, while the *Sant’Ambrogio* hospital and the *Ospedale della Pietà* respectively 18 and 11. These data confirm that in 1480 a large network of hospitals was active in Milan. Among them, the Ospedale Maggiore, strongly desired by Francesco Sforza, whose first stone was laid on April 12, 1456, and which had begun to function in 1472, while it was still under construction, had not yet replaced the small hospitals, assuming the predominant role in health assistance to which it was intended by the duke. Not considering the deaths in the foundling hospital, the deaths in hospitals, therefore, represent 0,07% of the deaths recorded in Milan in 1480.

## Final considerations

The Mortuorum Liber of Milan is almost unique, compared to the other European register of deaths, both for the precocity of its establishment and the richness and completeness of the information contained. Directly promoted and managed by the ducal government and based on the direct inspection of each corpse by a public official, and a rigorous data collection, the system organized by the Sforza is of extraordinary modernity. The methods of certifying deaths still in use in Italy^16^ and in advanced health systems are largely indebted to and in many respects still similar to the Sforza system, whose main limitation is the non-publicity of data, which remained handwritten and can only be consulted by health authorities for centuries.

The Mortuorum Liber of Milan was established to manage the potential presence of plague in the city, in general, during an epidemic event, it is important to identify the cases rapidly to put in place measures to contain the contagion. This is also important for modern epidemics, the recent one is the COVID-19, to the extent that reporting delay can cause the distortion between the real incidence of the disease and the reported one (Sarnaglia et al. 2021). The incidence and the prevalence are important to take measures to protect the population.

As a part of the Sforza’s health project addressed to preventing plague, the MiSfoRe implied a foresightful vision of the community as an interdependent whole, beyond class and economic distinction, with a death registration program based on the personal responsibility of public officials in the collection of information at community-level and the cooperation of the population. The sharing of risk events joins dead and ill people with civil servants and the community, through a common engagement of the body which underlies a wider vision of corporeality as heritage and transmission. It would be not too ambitious to envisage the Milano Sforza Registers as the first model to monitor the health landscape of the city by the inclusion of people of all ages and all social conditions through a detailed report of information. The result, especially for the 15^th^ century, is a wealth of precious data that deserve to be made available to the scientific community.

The fruition of the information contained in the registers by scholars of different disciplines – very limited to date -could be made possible by the digitization project of the data contained in the registers that we intend to carry out. Analysis of the deaths of 1480 reported in this paper, which in our opinion represents a not insignificant contribution to the knowledge of the conditions of life and of dying in a Renaissance city. In particular, despite limited attention being paid to the causes of death in childhood, the data on children’s deaths contained in the MiSfoRe are however of strong interest and broader than those of other ancient registers, where they are often completely absent. Perinatal death is the single most frequently-cited ’cause’ of death, even if without any further specification of the pathological events associated with it. Similarly, in the large majority of deaths that occurred in children, no cause specification was available. However, studies on the distribution of deaths in children and the general population in the different areas of the city may represent the basis of future studies addressed to estimate the total population in the city as so as in each parish or city area and to assess the socio-economic differences within and among them.

The picture of Milan that the register of 1480 gives us paints a city whose interest in caring for the sick is testified by the many functioning hospitals, in which, however, died only a relatively small percentage of the citizens died during the year. A Milan in which the high number of deaths reported in children suggests high infant mortality, having a decisive effect on life expectancy, without however preventing a non-negligible number of citizens from reaching - or presuming to have reached - one hundred years of age. A city in which deaths from hunger, although not absent, seem to be very few. A city with a non-negligible number of murders, which however seem to concern only men who have faced each other in white weapon fights.

In conclusion, the MiSfoRe not only illustrates the historical landscape of a sanitary policy in the early Renaissance but also illuminates an extraordinary adherence between ethics and method through a strict and meticulous division of tasks, which could be considered as an embryonal protocol of professional deontology. Above all, they offer an exceptional basis for further studies involving multiple disciplines. For this purpose, also to guarantee data preservation, the MiSfoRe digitization is desirable and necessary.

## Data Availability

The data are available from the original source on the Middle-Age registers at Archivio di Stato di Milano, Italy

## Acknowledgements

The Authors are grateful to Dr.Benedetto Luigi Compagnoni, Director of Archivio di Stato di Milano for letting access the *Mortuorum Libri*

## Footnotes

1 In such a period, so frequently tormented by the recurrence of the epidemics, just to give an example more than 35 treatises on plague have been published in Italy including the *De peste,* wrote in 1478 by Jacopo Soldi; the *Trattato de la pestilentia* by Girolamo Manfredi (1479), the *Consilium contra pestilentia* by Gentile da Foligno (1479), and *De Pestilentia* by Giovanni Calori. In 1481 Marsilio Ficino published the *Consilio contro la pestilentia*, which was republished several times between 1575 and 1580 when strong plague epidemics resurfaced (Katinis 2007) (Manfredi 2008).

2 Registers were written in Latin before 1774

3 Data referring to social status and employment are largely incomplete and reported only in some cases, especially for ordinary people. The registers that most accurately report this kind of data are those of the years from 1485 to 1495. (Zanetti 1976)

4 (1.339.000, with 51 cities) with respect to France (688.000, 32 cities), Spain (414.000, 20 cities), Germany (385.000, 23 cities), Belgium (295.000, 12 cities), The Netherlands (150.000, 11 cities) and England and Wales (80.000, 5 cities) (Malanima 1998). In Italy, the largest city was Venice, with 150.000 inhabitants estimated in 1423 (Spruyt 1996), while Florence had 54.000 inhabitants in 1470 (Chandler 1987) and London about 50.000 in 1500 (Chandler 1987)

5 The possible fatal outcome of the reported case was also reported, writing “ M ° ” for mortal ones. that is“ mortal ”. The payment of the burial tax, if due and paid“ S ”(*solvit*),“ 0 ”if not due, such as for infants, or“ I ” for insolvent. (Folco Vaglienti, personal communication).

6 In 1468 there were two deputies with authority over the entire ducal territory, Pietro Trivulzio and Franceschino di Castelsampietro, drawn respectively from the Secret Council and the Magistracy of Extraordinary Revenue, two of the highest institutional organizations of the Sforza central government (Vaglienti 2020a; Vaglienti 2020b; Gamberini 2020).

7 The deaths attributed to specific diseases will be illustrated in further publications.

8 The data entry was performed by the staff collaborating with the paleographer.

9 A map of Pietro De Guidoldi, designed at the end of the 14^th^ century, shows the roman and the medieval rings as two concentric circles, not responding to reality but according to the concept that the circle represents perfection. However, he reported on the map precise details about the name of waterways and some buildings and infrastructures. A second map of the same Author has the *Broletto* in the center. In a perspective map of Milan, dated back to the end of 1400 and the beginning of 1500, the walls of Milan were also represented with round shapes, with the Dome at this center and sketches of buildings along the ways conducting to walls. Canals were also represented. In a map contained in the *Codice Atlantico* of Leonardo Da Vinci, the square in the middle represents the “real” center of Milan: the ancient Roman forum, where the cardo and the decumanus joined (Navoni 2018). Despite even in Leonardo’s sketch the city is drawn with a roughly circular shape, it is interesting that he placed the castle in its real position, at the northwest limit of the city, while the previously cited maps and subsequently many others, up to the beginning of the 17^th^ century, placed it at the top of the circle in which the city was ideally contained.

10 “The children, after being welcomed, were baptized - if there was no certainty that the sacrament had already been administered - registered and then entrusted to the internal nurses for breastfeeding” (P. 40). From 18 months to four years they were assigned to the nurtures, after this period they re-entered to receive an education (lay and religious) and to receive professional formation (male as artisans and female in embroidery and ribbons) (Canella 2009).

11 “*Si come dice l’autoritade’ de’ fisici che sono tre generazioni di febbre; effimera, e etica, e putrida. E la ragione perchè sono solamente tre generazioni, si puote sapere per l’autoritade di Isaac nel libro delle febbri, che dice che il corpo umano è composto di tre modi; cioè spiriti, ossa, e umori. E nelli spiriti si genera effimera, e nell’ossa etica, e nelli umori putrida.* (As the authority of physicians says, there are three generations of fever; ephemeral, ethical, and putrid. And the reason why they are only three generations can be known by Isaac’s authority in the book of fevers, which says that the human body is composed of three modes; that is, spirits, bones, and humors. And in the spirits ephemeral is generated, and in the bones ethics, and putrid in the humors)”. Anonymous manual of the 14^th^ century, edition by G. Manuzzi, Florence 1863, page 1. According to Avicenna, ethical fever is a radical fever of the limbs, which disturbs the spirits by heating them and evaporating the humidity.

12 as exemplified by the unfortunate case of Bertolino del Signore, killed at 26 a letali vulneri in radice sinistre partis colli (by a fatal wound at the base of the left side of the neck) or by the case of Francesco da Desio, who died at 24 after “hit with a sword on the skull and also near the nostrils, that show the deep signs, with intervened fever, after the 20^th^ day”.

13 *Semifatua nec ex toto sui iuris, se ipsa et de mente propria arsenicho venenavit et yta proprio presbitero confessa est penitentiam agens et in presentia Johannis Petri de Affori et Bernardini speciarii regis et sui famuli et Josep de Ferrariis quibus fides adhibenda est, presentibus etiam duabus matronis fidedignis* (Semidiot and not totally in the full capacity of her faculties, she poisoned herself with arsenic and on her initiative and therefore she confessed to her presbyter repenting and in the presence of Gio.Pietro da Affori and Bernardino, the sovereign’s apothecary, and his servant and Giuseppe Ferrari, to whom credit must be given, also present two trustworthy married women).

14 *Studiose animi, incitata passione, assumpto veneno argento sublimato die martis proxime passata colirica et mortali succedente passione* (With premeditation, driven by passion of love, taken for poison mercury last Tuesday, with an ensuing fatal colic attack).

15 In the present time the declaration of death can be done only by a doctor, in particular the *Necroscopo*, if the event occurred outside a medical structure, who draws up the death certificate. The doctor who cares for the deceased (even if he did not directly witness the death) has the task to write the ISTAT (*Istituto Nazionale di Statistica,* National Institute of Statistics) form, which is important for an epidemiological and statistical point of view. It consists in a sanitary part and an anagraphic one (Blog MMG Medicina Generale 2017). The sanitary part is compiled by the doctor and contains: gender, place and the territory of the event, sequence of conditions that led to death and time elapsed between the onset of the cause and death. If the death is due to a traumatic event or poisoning, the mention of modality is requested (accidental, homicide or suicide). Other information are requested incase of transport accident. The anagraphical part, completed by the civil status office, contains: gender, date of the event, the location of the birth, information about the profession of the deceased, age, marital status, residence and citizenship (ISTAT 2020).

16 The case of this elderly gentleman, who belonged to an aristocratic family included in the *Matricula nobilium familiarum Mediolani* with the act dated 20 April 1377, describes a typical case of death in an elderly person following a fracture of the femur. The *Necroscopo* verify the congruence between the ISTAT form and the results of his visit (Blog MMG Medicina Generale 2017). In addition the certification of the causes of death was of competence of doctors both in the past and now for the city of Milan. These elements reveal similarities between the information contained in the Registers of Milan and the modern ISTAT form. Consequently it would be not too ambitious to envisage the Milano Sforza Registers as the first model to monitor the health landscape of the city by the inclusion of people of all age and all social conditions through a detailed report of information.

